# Describing the landscape of nutrition or diet-related randomised controlled trials: meta-research study of protocols published between 2012 and 2022

**DOI:** 10.1101/2023.06.08.23291109

**Authors:** Flávia Moraes Silva, Amanda Rodrigues Amorim Adegboye, Cintia Curioni, Fabio Gomes, Gary S Collins, Gilberto Kac, Jonathan Cook, Leila Cheikh Ismail, Matthew J Page, Neha Khandpur, Sarah Sallie Lamb, Sally Hopewell, Shaima Saleh, Shona Kirtley, Simone Bernardes, Solange Durão, Colby J Vorland, Michael Schlussel

## Abstract

**Background & Aims:** The landscape of nutrition research has changed over the past decades. We aimed to map the landscape of nutrition or diet-related interventions research, using data from randomised controlled trial (RCT) protocols published in the last decade.

**Methods:** This meta-research study examined nutrition or diet-related RCT protocols published in journals indexed in PubMed, Embase, Cinahl, Web of Science, PsycINFO, or the Global Health Database between January/2012 and March/2022. Two reviewers independently screened the titles and abstracts to check eligibility and one reviewer extracted bibliometric information, study characteristics, and research transparency practices such as protocol registration, conflicts of interest and funding disclosure. We also screened the "Instructions for Authors" of journals with publications in our sample to check for endorsement of SPIRIT, TIDieR, and CONSORT reporting guidelines, and we checked if the authors mentioned these reporting guidelines in their paper.

**Results:** The search retrieved 62,319 records, of which 1,068 met the eligibility criteria and were included in the review. The number of published RCT protocols increased annually between 2012 and 2022, with a mean of 161 (range: 155-163) publications/ year. The USA (n = 165; 15.5%) and Australia (n = 137; 12.8%) published the largest number of protocols. Protocols were published in 148 journals, mainly medical journals (n = 518; 48.5%). Among these journals, 50 (33.8%) endorsed SPIRIT, 111 (75.3%) endorsed CONSORT, and four (2.7%) endorsed TIDieR. In 343 (32.1%) publications the authors mentioned SPIRIT, in 297 (27.8%) CONSORT was mentioned, while 20 (1.9%) mentioned TIDieR. Most protocols reported the RCT registration number (n = 1,006; 94.2%) and included statements about conflicts of interest (n = 952; 89.1%) and funding (n = 994; 93.2%). More than one third of protocols focused on adults and elderly participants (n = 350; 32.7%) and most protocols included participants with a specific clinical condition (n = 726; 68.0%). A single nutrition or diet-related intervention (n = 724; 67.8%) was described in most protocols, with "supplementation, supplements or fortification" (n = 405; 37.9%) and "nutrition education, counseling or coordination of care" (n = 354; 33.1%) being the most frequent types of interventions studied. The most frequent primary outcomes reported were related to clinical status (n = 308; 28.8%), nutritional status (n = 247; 23.1%), and frequency or severity of disease (n = 238; 22.3%). The majority of protocols described a single-centre study (n = 838; 78.5%), with two-arms (n = 844; 79.1%), parallel (n = 1014; 94.9%) design, with a superiority framework (n = 755; 70.7%).

**Conclusions:** The number of protocols on nutrition or diet-related trials being published is increasing, indicating the importance of this type of publication. The mention of relevant reporting guidelines by both researchers and journals remains far from ideal. Most protocols assessed supplementation or fortification and nutrition education, counselling or coordination of care interventions, among adults and the elderly.

## Introduction

Nutrition research aims to provide the evidence basis for establishing dietary guidelines and also support both clinical practice and public health recommendations and policy making. To keep advancing the field, and ensure that nutrition scientists achieve its main goal of improving the general population’s quality of life and patient care, the conduct and reporting of research should be of high-quality (1,2).

Randomised controlled trials (RCTs) provide important evidence for clinical decision-making (3). Making RCT protocols publicly available has been supported as good research practice over the last decades since it contributes to increase research transparency and rigour (4). Besides registration of RCTs being mandatory by several research sponsors, funders, and journals in several countries, as well as recommended by the International Committee of Medical Journals (5), making the study protocol publicly available has the advantage of providing a more complete and detailed description of the planned research, in comparison to the limited templates offered by registration platforms (4, 6–8).

Publicly available RCT protocols help to ensure consistency of trial procedures, ethical assumptions, transparency, and reliability of research findings (8). Having a protocol submitted as a scientific article early in the research pipeline potentially increases research quality, as it provides researchers with the opportunity for considering the opinions of external experts, aids with the interpretation of study results, and reduces selective outcome reporting (7,8).

The concerns related to the quality and integrity of research published in the field of nutrition reflect those observed for other fields and recognised as a "credibility crisis" (9). In response to this crisis, the scientific community has called for more rigour and transparency in the editorial process of scientific journals (10,11), including specific requests for detailed statements on conflicts of interests and funding, preregistration of hypothesis and study methods, and the endorsement of reporting guidelines (11). However, the adoption of such practices remains far from optimal. In several biomedical disciplines, including nutrition, less than 50% of journals endorse reporting guidelines (11,12).

As far as we know, the frequency of publishing protocols of nutrition or diet-related RCTs as well as the scope and methods described in such publications, has not been previously investigated. We therefore aimed to map the contemporary landscape of nutrition or diet-related interventions research, based on RCT protocols published between 2012 and 2022. We also aimed to investigate the adoption of research transparency and reproducibility practices in these publications.

## Methods

This

### Study design and eligibility criteria

We extracted data from protocols of nutrition or diet-related RCTs published as scientific articles (registration: https://doi.org/10.17605/OSF.IO/YWEVS). Full details of the protocol can be found elsewhere (13).

We searched for relevant protocols on PubMed, Embase, Cinahl, Web of Science, PsycINFO, or the Global Health Database between 01/January/2012 and 24/March/2022. Study design was initially assessed based on self-identification by the trialists (i.e., whether the authors described their studies as RCTs). We applied no restriction to the population or outcomes studied, or to the publication language.

Nutritional interventions combined with others (such as exercise or drugs), or as part of a lifestyle or health program intervention, were eligible. The following types of intervention were included: a) diets, dietary components, and dietary patterns; b) formulated, fortified, and enriched foods; c) dietary products, including dietary supplements; d) nutrients and bioactive non-nutrient components naturally present in foods (e.g., cinnamon); and e) nutritional education, promotion, counselling, or programs (14). We excluded protocols of RCTs that only used pharmaceutical or herbal medicines as nutrition intervention, protocols of non-randomised trials, and publications reporting the study findings.

### Literature search

The lead author (FMS) and a professional health sciences information specialist (SK) built a search strategy for PubMed (via the National Library of Medicine) combining the search strategy developed by Durão et al. to identify diet and nutrition trials (15) and a modified version of the search strategy developed by Madden et al. to identify RCT protocols (16). We adapted the search strategy to Embase (via Elsevier), Cinahl (via EBSCO), Web of Science (via Clarivate), PsycINFO (via Ovid), and Global Health Database (via Ovid). On March 24, 2022, we ran the search strategies for all databases (see complete search strategy in the Supplementary Box 1).

### Selection of eligible reports

We imported all retrieved references into EndNote^®^ and used its automated deduplication feature to remove duplicates. We exported the records to Rayyan^®^ (17) and the lead author (FMS) manually double-checked the resulting reference list and removed any remaining duplicates. Two reviewers (FMS and JL) then independently screened the publications’ titles and abstracts to check for eligibility, followed by screening of potentially eligible full-texts by one author (FMS). Disagreements between reviewers were resolved by consensus.

### Data collection

One reviewer extracted data using a standardised data extraction form designed on REDCap® (18). Data were cross-checked in a sample of 100 protocols by another reviewer (SS) with a concordance rate of 96.5%, ranging from 89.1% to 100%.

Information collected from included protocols were the first author’s name, journal and year of publication, bibliometric information, research transparency practices (e.g., funding and conflicts of interest statements (yes/no), and details of protocol registration), and general study characteristics (information about the participants, interventions, comparators, outcomes, and study designs). **Box 1** describes the nutrition intervention categories of interest, which were adapted from Naude et al. (14).

**Box 1:** PICOS categories adopted for data extraction of diet and nutrition-related trials’ protocols.

| Categories used for data extraction |  |  |
| --- | --- | --- |
| **Participants** | Pregnant women<br>Infants<br>Adults<br>Adults and elderly<br>Postmenopausal women | Mother and infant pairs<br>Children and preschool-aged children<br>Elderly<br>Families<br>Participants with a clinical condition |
| **Interventions<sup>†</sup>** | Food (whole food, food products, specially formulated foods)<br>Breastfeeding, complementary feeding, weaning<br>Complete diet or dietary pattern<br>Complete nutrition formulas (enteral or parenteral)<br>Supplementation, or supplements, or fortification (single or multiple nutrients, bioactive non-nutrients, plant components)<br>Nutrition education, counselling, and coordination of care<br>Other, if no component of intervention could be categorised as any of the above |  |
| **Comparator** | Placebo<br>Usual care<br>Other | No intervention<br>Different intervention |
| **Outcomes** | Mortality<br>Clinical status (clinical or biochemical measures)<br>Nutritional status (anthropometry, body composition, nutrition diagnosis)<br>Frequency or severity of disease<br>Diet quality and/or variety<br>Food/ nutrient/ dietary intake<br>Diet-related behaviours<br>Other non-dietary behaviours<br>Withdrawal from the study, drop-out or adherence-related<br>Adverse events, side-effects and/or safety<br>Cost-effectiveness or economic<br>Quality of life<br>Other |  |
| **Study design** | Parallel<br>Cluster<br>Pilot | Crossover<br>Factorial<br>Multicenter |
| **Study framework** | Superiority<br>Non-inferiority<br>Not reported | Equivalence<br>Exploratory |

<sup>†</sup>Adapted from Naude et al. (13)

One reviewer (SB) collected data on journals’ endorsement of the following reporting guidelines: SPIRIT (8), CONSORT (19), and TIDieR (20). The "Instructions for Authors" webpages of each journal identified in our sample were screened, and endorsement was characterised by the presence of either a specific requirement that authors should adhere to the relevant reporting guidelines’ checklists when writing their manuscripts (regardless of whether the complete checklists should be submitted or not) or any general recommendation to follow these reporting guidelines. One reviewer (FMS) also checked if the authors mentioned these reporting guidelines in their paper (i.e., self-reported adherence to reporting guidelines or formal citation).

### Amendments to the protocol

In addition to the questions included in the original data extraction form (13), we added the following questions: 1) Is it a pilot study? (yes, no); 2) What is the framework of the RCT? (superiority, equivalence, non-inferiority, exploratory, not reported); 3) Country where the RCT is being planned; 4) Was the RCT registered? (yes, no) If so, where? (registration platform name); 5) Detailed description of the intervention; 6) Intervention duration (in days), if delivered for a fixed period; 7) Declaration of conflicts of interest; 8) Funding statement. The final data extraction form is available as supplementary material.

We also decided to evaluate endorsement of the reporting guidelines by all journals in which the protocols were published, as we noticed a low prevalence of citations to these documents in the included publications. We also collected the 2021 impact factor of all journals through the Web of Science database.

### Data analysis

Journals were grouped into three categories according to the scientific research field: medical or health-related, methods, and nutrition journals. The clinical conditions of participants were grouped according to the types of diseases. Malnutrition and critically ill patients were also included as categories for describing the clinical condition of participants. Subgroup analyses according to the diagnosis of cancer (present or absent) and cardiovascular diseases were conducted, since these are the major causes of death globally (21).

The intervention categories ‘complete diet or dietary pattern’ and ‘supplementation, supplements or fortification’ details were grouped according to the type of diet and supplements, respectively. Protocols were grouped according to the duration of the intervention into three categories: fixed (if the authors described a unique period for all participants), not fixed (if the duration of the intervention depends on the incidence of outcomes and it is not the same for all participants), and not reported. Interventions were also classified as ‘of acute response’ if the outcomes would be evaluated 24 hours after delivering the intervention. More information can be found in the data analysis section of the Supplementary Material.

We explored the frequency of each component of PICOS (participants, intervention, comparator, outcomes, study design) and practices of transparency and reproducibility among protocols according to the year of publication, the countries where the trials would be conducted (the five most frequent), and the subgroups of protocols involving patients with cancer and cardiovascular diseases, to investigate if these features could explain some difference in the nutrition or diet-related RCT protocols.

The statistical package SPSS 22.0^®^ was used for data tabulation and analyses. We calculated the absolute and relative frequency of all categorical variables and presented the results as n (%). For quantitative variables, medians and ranges (minimum-maximum) are presented. Graphics were designed in Excel®.

## Results

### Literature search and protocols selection

The literature search retrieved 62,319 records, and the titles and abstracts of 40,389 deduplicated records were screened. Of these, the full-texts of 1,192 records were screened, with 121 publications identified as ineligible. Protocols of 1,068 RCTs met inclusion criteria and were included in this meta-research study. The detailed selection process of nutrition or diet-related RCT protocols is presented in **Figure 1**.

**Figure 1.**
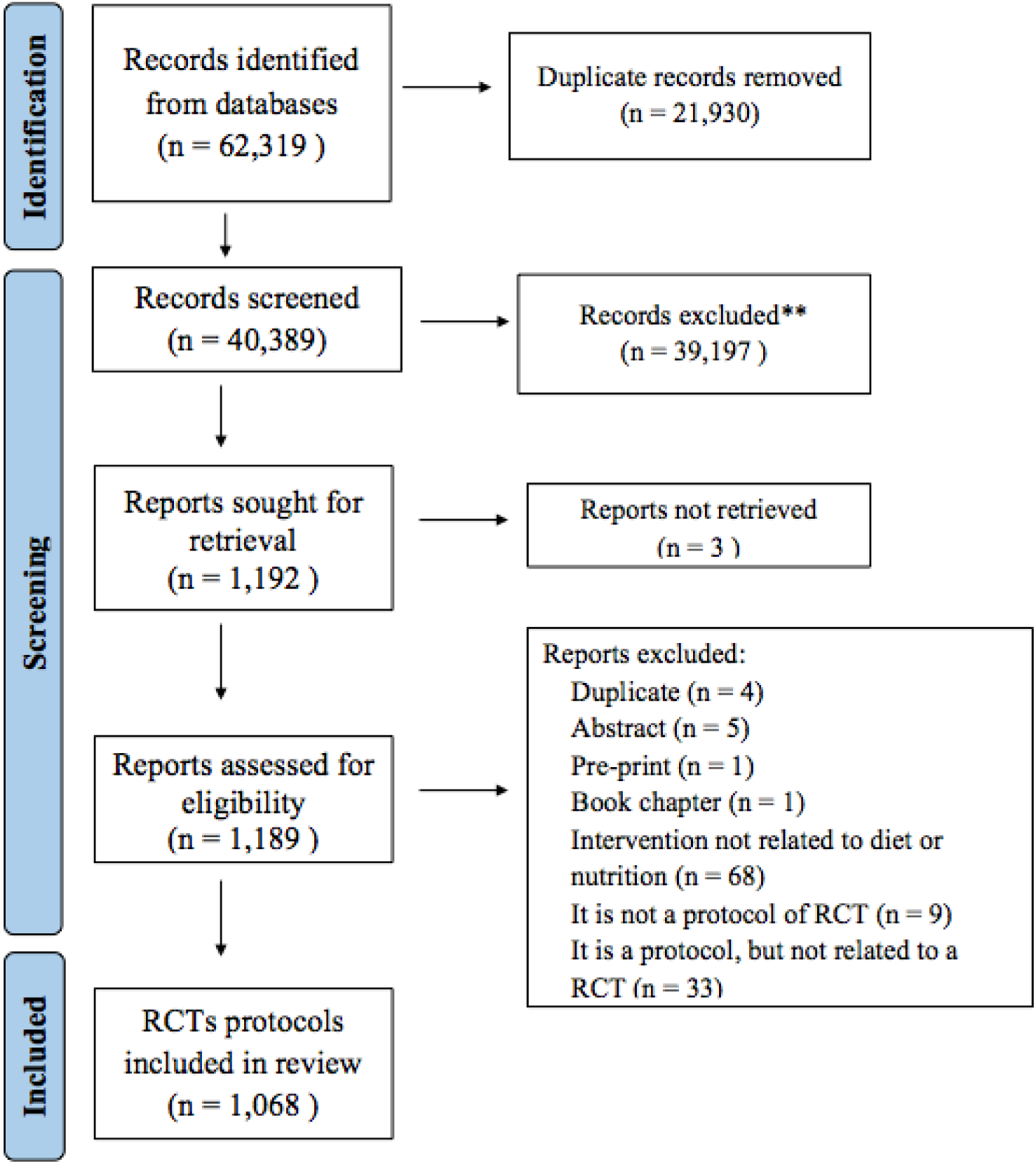
Flow chart of nutrition or diet related RCT protocols selection. RCT = randomised controlled trials.

### General characteristics of the publications

The number of published nutrition or diet-related RCT protocols has increased annually between 2012 and 2021, as demonstrated in **Figure 2**, with a mean of 161 (range: 155-163) publications/year, in the last three years of the period. The countries publishing the largest number of protocols are the USA (n=165; 15.5%), Australia (n=137; 12.8%), UK (n=72; 6.8%), Iran (n=65; 6.1%), and China (n=65; 6.1%) **(Figure 3).**

**Figure 2.**
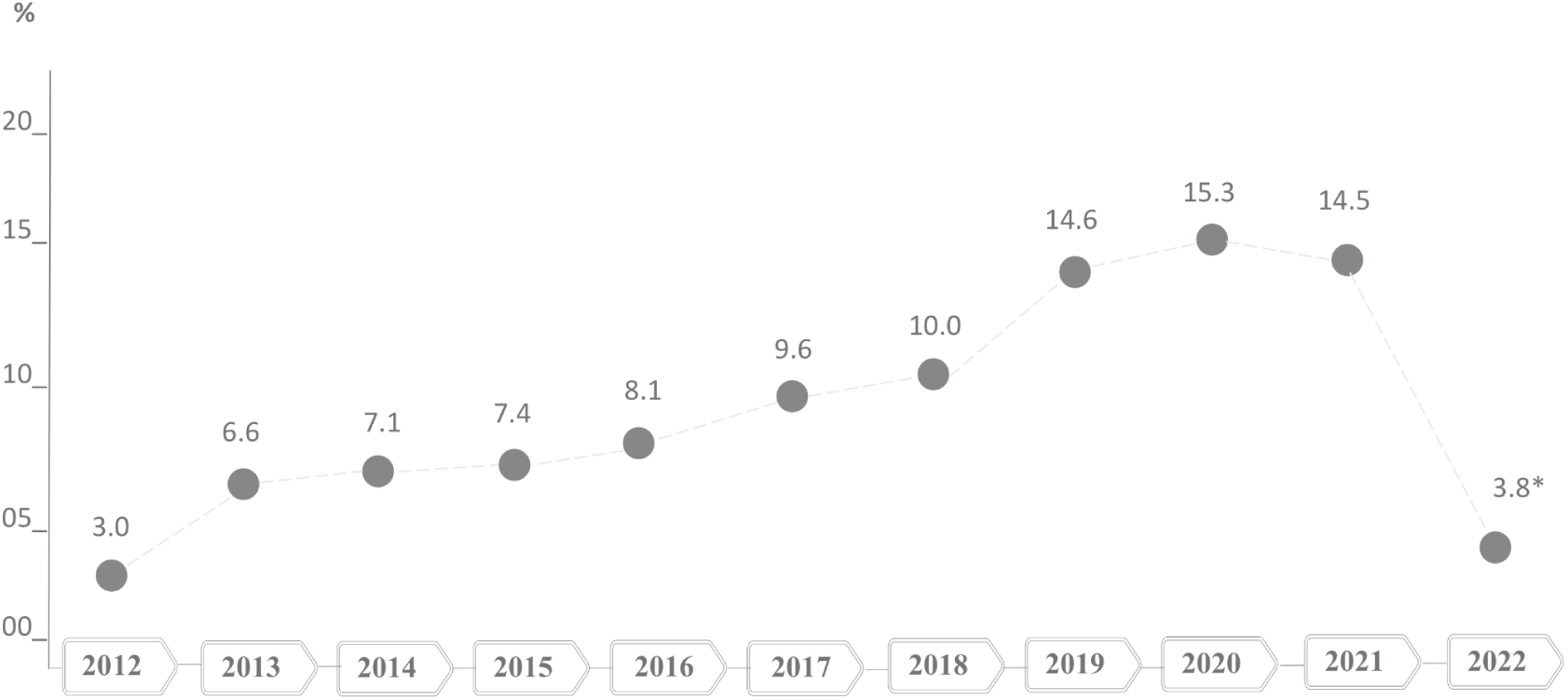
Relative frequency of nutrition or diet related RCT protocols published between 2012 and 2022 (* corresponding to the first three months of 2022).

**Figure 3.**
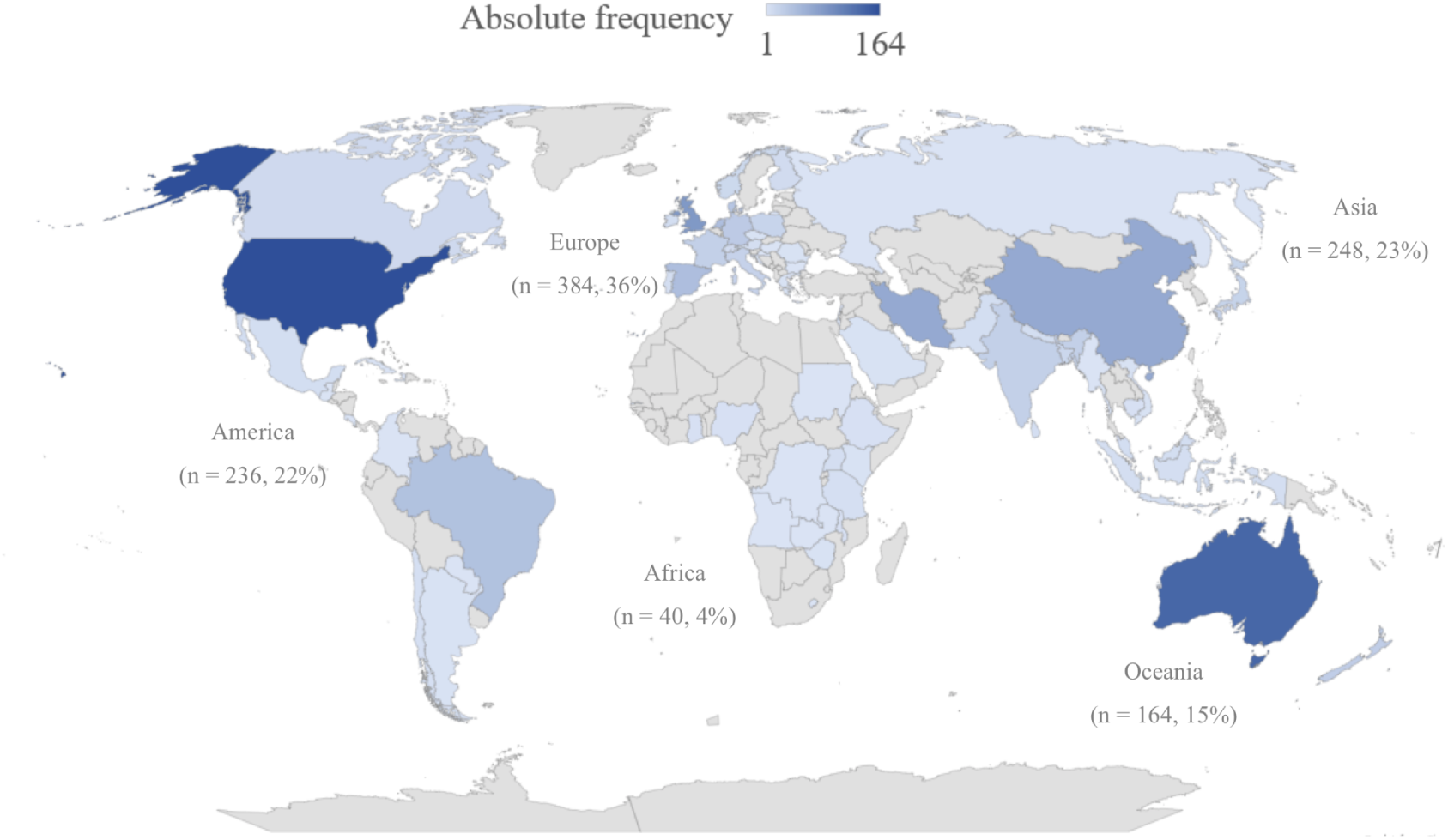
Distribution of published nutrition or diet related RCT protocols between 2012 and 2022 in the world. Data are shown as absolute frequency by countries and absolute (relative) frequencies by continent.

Most protocols (n=1,006; 94.2%) reported that they were registered and “ClinicalTrials.gov” (n=520; 48.7%) was the most used registration platform, followed by the Australian and New Zealand Clinical Trial Register (n=154; 14.4%), and the International Standard Randomized Clinical Trial (n=117; 11.0%). Most publications included a statement about conflicts of interest (n=952; 89.1%), and among these 783 (82.3%) declared no conflicts of interest. More than 90% of the publications included a funding statement (n=994; 93.2%). Only 48 (4.5%) publications declared that the RCT was not funded.

The majority of nutrition or diet-related RCT protocols were published in medical journals (n=518; 48.5%), followed by methods journals (n=479; 44.9%), and nutrition journals (n=71; 6.6%). A total of 148 journals published these protocols, and the journals with the highest number of protocols published between 2012 and 2022 were Trials (n=295; 27.6%), BMJ Open (n=153; 14.3%) and Contemporary Clinical Trials (n=83; 7.8%) (**Table 1**). For 44 (2.7%) journals the impact factor was unavailable, while for the remaining ones, it ranged from 0.813 to 20.999.

**Table 1:**
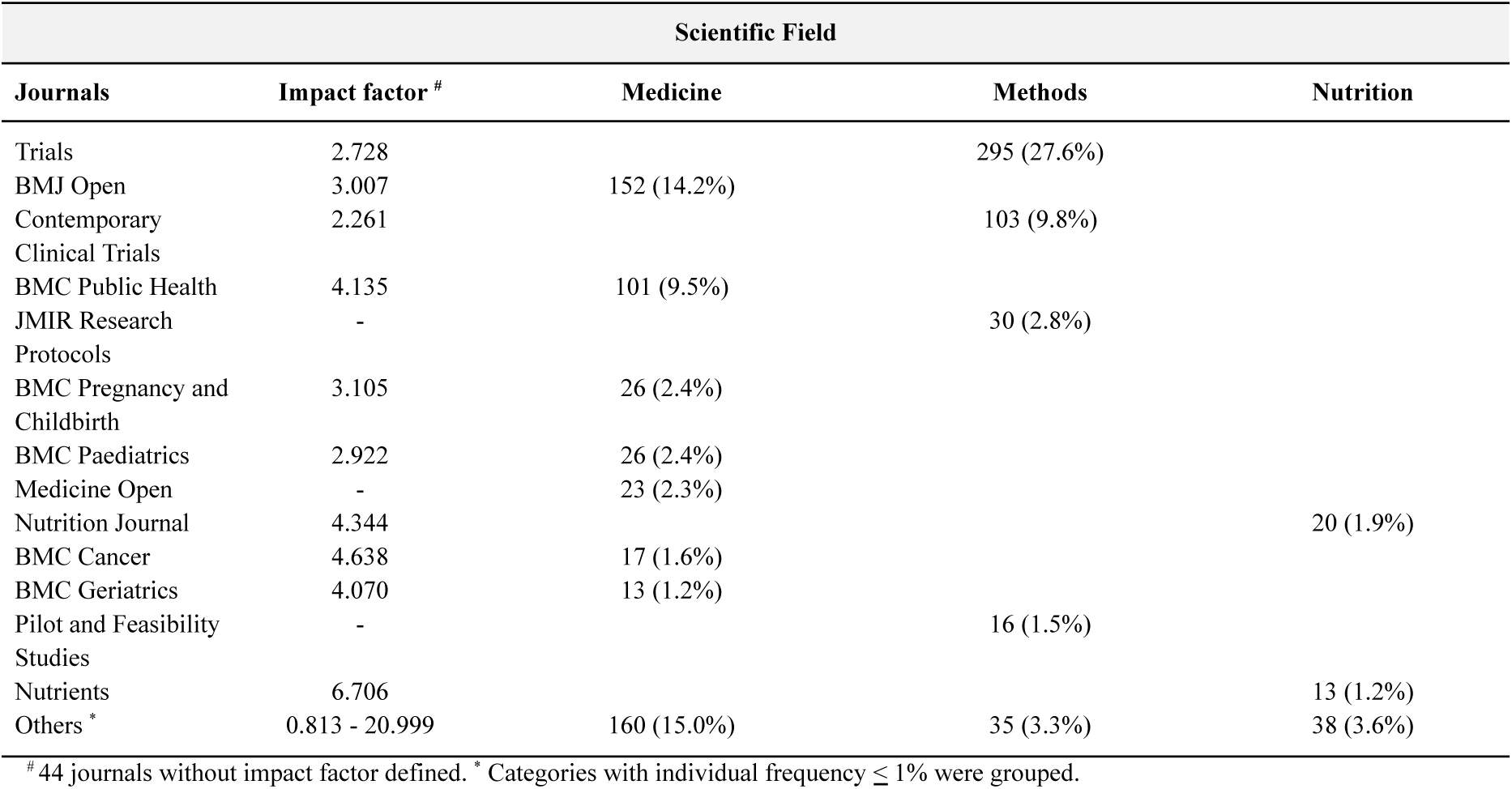
Frequency of published protocols of nutrition or diet-related RCTs according to the journal’s scientific field categories.

| Scientific Field |  |  |  |  |
| --- | --- | --- | --- | --- |
| Journals | Impact factor <sup>#</sup> | Medicine | Methods | Nutrition |
| Trials | 2.728 |  | 295 (27.6%) |  |
| BMJ Open | 3.007 | 152 (14.2%) |  |  |
| Contemporary Clinical Trials | 2.261 |  | 103 (9.8%) |  |
| BMC Public Health | 4.135 | 101 (9.5%) |  |  |
| JMIR Research Protocols | - |  | 30 (2.8%) |  |
| BMC Pregnancy and Childbirth | 3.105 | 26 (2.4%) |  |  |
| BMC Paediatrics | 2.922 | 26 (2.4%) |  |  |
| Medicine Open | - | 23 (2.3%) |  |  |
| Nutrition Journal | 4.344 |  |  | 20 (1.9%) |
| BMC Cancer | 4.638 | 17 (1.6%) |  |  |
| BMC Geriatrics | 4.070 | 13 (1.2%) |  |  |
| Pilot and Feasibility Studies | - |  | 16 (1.5%) |  |
| Nutrients | 6.706 |  |  | 13 (1.2%) |
| Others <sup>*</sup> | 0.813 - 20.999 | 160 (15.0%) | 35 (3.3%) | 38 (3.6%) |

<sup>#</sup> 44 journals without impact factor defined. <sup>\*</sup> Categories with individual frequency $\leq 1\%$ were grouped.

Of the 148 journals that published the included protocols, 50 (33.8%) endorsed SPIRIT, 111 (75.3%) endorsed CONSORT, and four (2.7%) endorsed TIDieR, with one of these explicitly endorsing its use for the reporting of protocols. While 95 (64.2%) of these journals had an unspecific endorsement of reporting guidelines (characterised by recommending searching for reporting guidelines on the EQUATOR Network website), 54 (36.5%) had a formal requirement for submitting the relevant reporting guideline’s checklist together with the manuscript, as observed on journals’ ‘instructions to authors’ webpages.

In 343 (32.1%) publications, the authors mentioned the SPIRIT guideline, in 297 (27.8%) they mentioned CONSORT, and only 20 (1.9%) mentioned TIDieR. The proportion of protocols mentioning CONSORT ranged between 18.8% (2012) and 35.4% (2015) during the studied period. The proportion of protocols mentioning TIDieR during the studied period varied from 0% from 2012 to 2014 (the year of its publication) to 3.2% in 2021, with a peak of 4.5% in 2019 (**Figure 4)**. Additional results can be found in **Supplementary Figure 1** and **Supplementary Table 1**.

**Figure 4.**
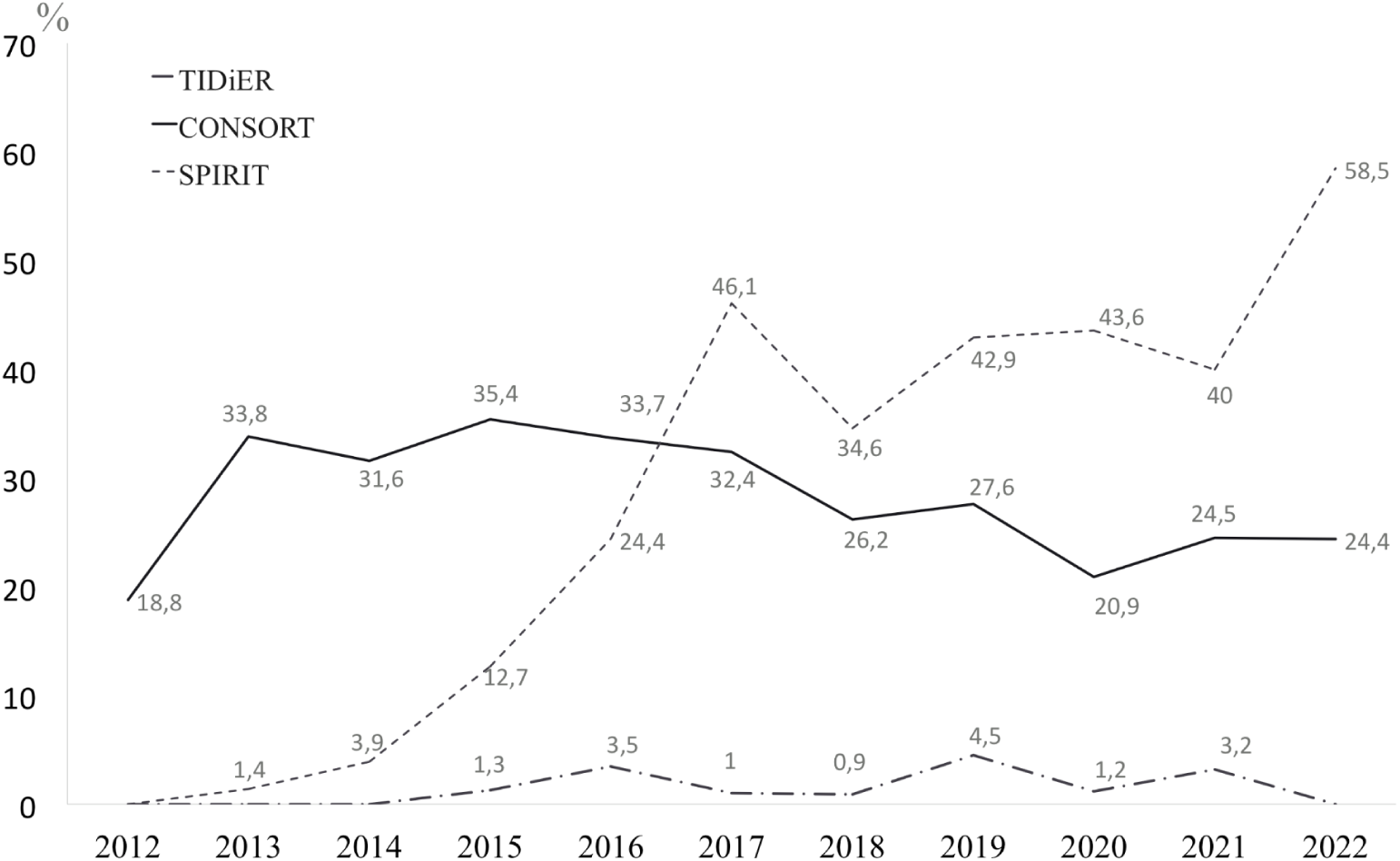
Relative frequency of nutrition or diet related RCT protocols that referenced SPIRIT, CONSORT, and TIDieR between 2012 and 2022.

### PICOS components of the nutrition or diet-related RCT protocols

**Table 2** characterises the protocols of nutrition or diet-related RCTs’ scope according to their PICOS. Most protocols described adults and elderly (n=350; 32.7%) or only adults (n=252; 23.6%) as the target population. In about one third of the protocols the target population was healthy individuals (n=342; 32.0%), while 21.5% included participants with endocrine (n=229) diseases, and 8.4% included participants with cardiovascular diseases (n=90).

**Table 2.** Details of nutrition or diet-related RCT protocols according to the categories of the PICOS acronym.

| **P - Participants** |  |  |  |
| --- | --- | --- | --- |
| *Group studied* | *Number (%)* | *Clinical Condition* | *Number (%)* |
| Adults and elderly | 350 (32.7) | None (healthy participants) | 342 (32.0) |
| Adults | 252 (23.6) | Endocrine diseases | 229 (21.4) |
| Children and preschool-aged children | 132 (12.3) | Cardiovascular diseases | 90 (8.4) |
| Pregnant women | 99 (9.3) | Gastrointestinal/ liver diseases | 82 (7.7) |
| Elderly | 76 (7.1) | Muscle and/or skeletal/ rheumatologic diseases | 49 (4.6) |
| Infants | 66 (6.2) | Infectious diseases | 39 (3.7) |
| Adolescents | 63 (5.9) | Psychiatric diseases | 31 (2.9) |
| Families | 53 (5.3) | Critically ill patients | 28 (2.6) |
| Mother and infant pairs | 19 (1.8) | Kidney diseases | 27 (2.5) |
| Postmenopausal women | 9 (0.8) | Neurological diseases | 25 (2.3) |
|  |  | Malnutrition | 24 (2.2) |
|  |  | Gynaecological diseases | 23 (2.2) |
|  |  | Lung diseases | 20 (1.9) |
| | | Others (individual frequency $\leq 1.0\%$) | 59 (5.5%) |
| **I - Intervention** |  |  |  |
| *Type* | *Number (%)* | *Categories* | *Number (%)* |
| Isolated | 724 (67.8) | Supplementation | 405 (37.9) |
| Component of a lifestyle intervention | 127 (11.9) | Nutrition education | 354 (33.1) |
| Combined with exercise | 98 (9.2) | Complete diet or dietary pattern | 165 (15.4) |
| Component of a health programme | 83 (7.8) | Foods | 78 (7.3) |
| Combined with drugs | 18 (1.7) | Complete nutrition formulas | 37 (3.5) |
| Combined with other medical care | 18 (1.7) | Breastfeeding | 17 (1.6) |
|  |  | Other | 12 (1.1) |
| **C - Comparators** |  |  |  |
| *Type* | *Number (%)* | *Number of arms* | *Number (%)* |
| Placebo | 362 (33.9) | 2 | 844 (79.1) |
| Usual Care | 316 (29.6) | 3 | 134 (12.5) |
| Other intervention | 289 (27.1) | 4 | 75 (7.0) |
| No control | 101 (9.5) | 5 or more | 15 (1.4) |
| **O - Outcome** |  |  |  |
| *Primary Outcome* | *Number (%)* | *Primary outcome categories* |  |
| Specified | 1021 (95.6) | Clinical status | 308 (28.8) |
| Not specified | 47 (4.4) | Nutritional status | 247 (23.1) |
|  |  | Frequency or severity of disease | 238 (22.3) |
|  |  | Food/ nutrient/ dietary intake | 68 (6.4) |
|  |  | Functional status | 57 (5.3) |
|  |  | Withdrawal from the study, drop-out or adherence | 56 (5.2) |
|  |  | Other non-dietary behaviours | 44 (4.1) |
|  |  | Diet-related behaviours | 39 (3.7) |
|  |  | Mortality | 35 (3.3) |
|  |  | Quality of life | 26 (2.4) |
|  |  | Breastfeeding | 19 (1.8) |
|  |  | Diet quality and/or variety | 17 (1.6) |
|  |  | Adverse events, side-effects and/or safety | 7 (0.7) |
|  |  | Cost-effectiveness or economic | 2 (0.2) |
|  |  | Other | 131 (12.3) |
| **S - Study design and Framework** |  |  |  |
| *Design* | *Number (%)* | *Framework* | *Number (%)* |
| Parallel | 1014 (94.9) | Superiority | 755 (70.7) |
| Crossover | 54 (5.1) | Exploratory | 92 (8.6) |
|  |  | Non-inferiority | 20 (1.9) |
| Cluster | 138 (12.9) | Equivalence | 7 (0.7) |
| Pilot | 72 (6.7) | Not reported | 194 (18.2) |
| Factorial | 39 (3.7) |  |  |

To estimate the effect of an isolated nutrition or diet-related intervention (n=724; 67.8%) was the most frequent aim of the included protocols. "Supplementation, supplements or fortification" (n=405; 37.9%) and "nutrition education, counselling or coordination care" (n=354; 33.1%) were the most frequent types of interventions studied. A total of 165 protocols (15.4%) aimed to evaluate the effect of a specific diet or dietary pattern. Only 101 (9.4%) protocols included no intervention (such as wait list) in the control group.

Among the protocols with "supplementation, supplements or fortification" as the intervention, vitamins (n=126; 31.1%), probiotics (n=63; 15.6%), and minerals (n=50; 12.3%) were the most frequent ones, as illustrated in **Supplementary Figure 2A**. The most frequent vitamins and minerals adopted as interventions were vitamin D (n=76; 60.3%) and iron (n=16; 32.0%), respectively. Among the protocols proposing to evaluate the effect of a specific diet or dietary pattern, the Mediterranean diet (n=26; 15.7%), Low-carb diet (including ketogenic or Palaeolithic diet) (n=22; 13.3%) and Energy-restricted diet (n=19; 11.5%) were the most frequently chosen, as demonstrated in **Supplementary Figure 2B**. A small number of protocols (n=38; 3.6%) planned to evaluate the acute response of an intervention (within 24h of exposure), while most protocols proposed to evaluate the response to the intervention after longer periods (n=899; 84.2%); with a median period under the intervention of 120 (minimum 2; maximum 2,160) days. In the remaining protocols (n=121; 11.3%) the period under the intervention was not fixed, meaning it was dependent on the incidence of the outcomes, or this information was not reported (n=10; 0.9%).

The most frequent primary outcomes reported by the protocols were "clinical status" (n=308; 28.8%), "nutritional status" (n=247; 23.1%), and "frequency or severity of disease" (n=238; 22.3%). Most protocols described a single centre study (n=838; 78.5%), with a two-arm (n=844; 79.1%), parallel (n=1014; 94.9%) design, and a superiority framework (n=755; 70.7%).

Characteristics of protocols according to the year of publication, countries, cancer and CVD diagnoses can be found in the Supplementary Results section.

## Discussion

This meta-research evaluated 1,068 protocols on nutrition or diet-related trials published in journals indexed in six online databases of medical literature between 2012 and 2022. The most frequent intervention applied was related to supplementation aiming to investigate its effects on indicators of clinical outcomes in adults and elderly with some disease. Most protocols were published in general Medical and health-related journals, and included the protocol registry, statements declaring conflicts of interest and funding source, though the minority of protocols mentioned a relevant reporting guideline.

Our results are consistent with those of a cross-sectional study on the scope and quality of Cochrane reviews of nutrition interventions published between 2007 and 2015, which also found supplementation to be the most frequently studied intervention (50%) and clinical or nutritional status assessment the most frequently evaluated primary outcomes (82.1%) (14). We did not observe a clear trend of these protocol details between 2012 and 2022. ’Supplementation, supplements, or fortification’ was the category of intervention most frequent between 2016-2018 and 2021-2022 whereas in the remaining years most protocols aimed to evaluate the effect of an intervention related to ’nutrition education, counselling and coordination care’. A potential explanation for ’supplementation, supplements, or fortification’ being the most frequent intervention planned in the reviewed protocols is that these are more feasible than a ’complete diet or dietary pattern’ or ’nutrition education, counselling, and coordination of care’ to deliver, as less behavioural modifications are required from the participants (14,22). Outcomes related to clinical status, nutritional status and frequency or severity of disease corresponded to more than 70% of all protocols, regardless of the year. This could reflect the fact that they can be achievable in the short-term considering that the medium of intervention duration was 120 days.

We also observed an increase in the number of protocols of nutrition or diet-related RCTs published as scientific articles, and most of these were registered in a clinical trial registration platform. Since the late 1990s, registration of such studies has been required by law in some countries (23). Good research practices include study registration because it can reduce publication and hindsight bias, safeguard honest research, and minimise research waste. A public registration record enables verification that the content of the research report corresponds to what was planned and described in the protocol (24–26). Furthermore, publishing RCT protocols as scientific articles contributes to increased transparency and robustness of research methods and findings (27,28). While some journals are increasingly supporting and publishing RCT protocols, this practice is still not common (29). A meta-research study of 326 RCTs found that only 36.2% of protocols were publicly available, with most of these available as peer-reviewed publications (47.5%) or as a supplementary file with the primary results (40.7%) (8).

We found that the minority of protocols in our sample were published in nutrition journals. This can be partially justified because protocols are not part of the scope of several nutrition journals, and accepting this type of publication has been a more recent practice among the journals that do so. Nevertheless, these findings highlight the importance of engaging with stakeholders from the wider scientific community, both to find a venue for such publications and ensure that nutrition or diet-related intervention research achieves its aims through methodologically robust and transparently reported studies. It also sends a clear message to editors of nutrition journals, as these are currently missing an important body of literature from their own area.

Greater transparency in disclosing all potential conflicts of interest can help stakeholders better understand what research questions are being proposed by whom, and the motivations behind such studies (30). An automated analysis of 2,751,420 open access records on PMC showed an upward trend in some reporting transparency indicators between 2000 and 2020, including conflicts of interest. For research articles, funding and conflicts of interest reporting percentages increased from 25% and 0% in 2000 to 89% and 91% in 2020, respectively (31). Despite the high prevalence of conflicts of interest and funding statements in the protocols studied, most of them were short and vague, providing little or no information about potential conflicts beyond financial ones. Besides financial conflicts of interest, indirect financial benefits, as well as non-financial conflicts of interest, can also influence research outcomes so should be disclosed.

Nutrition or diet-related trial protocols were published in 148 different journals, most of which endorse CONSORT, less than 35% endorse SPIRIT, and a minority endorse TIDieR in their ‘Instructions to Authors’. A meta-epidemiological study found that only 90 (53%) of 170 journals in the Endocrine and Internal Medicine area supported the CONSORT statement, with rates ranging from 9% (Hematology) to 63% (Internal Medicine), according to specialty (12). Another study examined the adoption of editorial procedures to improve the reporting of empirical studies in nutrition and dietetics research and showed that 27/30 (90%) of high-impact factor journals mentioned CONSORT, while 7/30 (23.3%) mentioned SPIRIT in the Instructions for Authors (11). These differences could possibly be attributed to the evolution of journals endorsement of reporting guidelines in the last decade, as well as the scientific field, since studies published in 2018 showed varying frequencies of CONSORT endorsement among the journals related to cardiology (5% of 19), critical care (14% of 37), dermatology (30% of 20) and oncology (52% of 21). Different methodology in the selection of the assessed journals in previous research can also explain these differences (12). Nevertheless, endorsement of reporting guidelines remains suboptimal, and journals can play an essential role in improving transparency in research reporting, as such endorsements indicate to authors the degree of completeness expected from them in their publications (32).

Obviously, journal endorsement of reporting guidelines does not guarantee adherence by authors. Thus, despite the positive increase in the number of protocols mentioning SPIRIT in the last years, that does not mean these protocols reported all the information required by its checklist. In the same sense, one cannot assume that a publication is not complete and transparent because a relevant reporting guideline was not mentioned in the text. Our next step is to assess reporting completeness in a subsample of the protocols described here, as part of a research programme to produce official developments for CONSORT, SPIRIT and PRISMA statements focusing on nutritional interventions (33). This is in line with the ongoing initiative of the Federation of European Societies to improve standards in the science of nutrition (34). We are in close contact with the FENS working group to gather expert input, increase dissemination of the final recommendations, and ensure a consistent message is presented.

We set up to describe the landscape of nutrition and diet-related interventions research, based on a sample of RCT protocols published in indexed medical journals. However, it should be kept in mind that, just like for any other interventions, the protocols of many RCTs in this area may never be published as articles (8). Yet, our sample of publications consisted of protocols published in journals indexed in six online databases of medical research, over a period of ten years, and our findings related to the main aspects of study scope, are similar to those previously described in the literature (14). Thus, we are reasonably confident that this work provides a good representation of the contemporary scenario of nutrition and diet-related intervention research. Another study limitation is the fact that we only performed a cross-sectional assessment of the current journals’ endorsement of reporting guidelines, which most likely have changed over the period in the scope of this study. Therefore, we might have missed important improvements in the endorsement of reporting guidelines by the journals in which the included protocols were published. Finally, future studies should explore more detailed information related to the disclosure of funding sources and conflicts of interest, since these can play a role in the transparency and reproducibility of nutrition or diet-related trials, particularly to understand whether these practices are related to the reporting completeness and risk of bias in RCTs and their protocols.

## Conclusion

The number of protocols on nutrition or diet-related RCTs being published is increasing, evidencing the importance of this type of publication. The adoption of relevant reporting guidelines by researchers at the design stage and their endorsement by journals remain far from ideal, potentially hampering the publication of RCT protocols as a mechanism of research transparency and integrity. Most protocols of nutrition or diet-related RCTs were not published in nutrition journals, underscoring the need to engage stakeholders beyond the nutrition research community to promote high-quality evidence generated by these studies and increase its impact. Our findings can be used by various stakeholders, such as researchers, institutions, funders, to assess what are the most commonly studied populations, interventions and outcomes in the field of nutritional interventions research, as well as the most frequent study designs employed to address these research questions and identify areas for future research focus.

## Data Availability

All data produced in the present study are available upon reasonable request to the authors

## References

1. Altman DG, Moher D. Declaration of transparency for each research article. BMJ. 2013; 347:f4796. doi: 10.1136/bmj.f4796.

2. Ioannidis JPA. The Challenge of Reforming Nutritional Epidemiologic Research. JAMA. 2018;320(10):969–970. doi: 10.1001/jama.2018.11025.

3. Zabor EC, Kaizer AM, Hobbs BP. Randomized Controlled Trials. Chest. 2020; 158(1S):S79–S87. doi: 10.1016/j.chest.2020.03.013.

4. Li T, Boutron I, Al-Shahi Salman R, Cobo E, Flemyng E, Grimshaw JM, Altman DG. Review and publication of protocol submissions to Trials - what have we learned in 10 years? Trials. 2016;18(1):34. doi: 10.1186/s13063-016-1743-0.

5. International Committee of Medical Journal Editors. Recommendations for the Conduct, Reporting, Editing, and Publication of Scholarly work in Medical Journals. Available on https://www.icmje.org/. Accessed on 22 May 2023.

6. Schönenberger CM, Griessbach A, Taji Heravi A, Gryaznov D, Gloy VL, Lohner S, et al. A meta-research study of randomized controlled trials found infrequent and delayed availability of protocols. J Clin Epidemiol. 2022;149:45–52. doi: 10.1016/j.jclinepi.2022.05.014.

7. Chan AW, Hrobjartsson A. Promoting public access to clinical trial protocols: challenges and recommendations. Trials. 2018;19(1):116. doi: 10.1186/s13063-018-2510-18.

8. Chan AW, Tetzlaff JM, Gotzsche PC, Altman DG, Mann H, Berlin JA, et al. SPIRIT 2013 explanation and elaboration: guidance for protocols of clinical trials. BMJ. 2013;346:e7586. doi: 10.1136/bmj.e7586.

9. Nuzzo R. How scientists fool themselves - and how they can stop. Nature. 2015;526(7572):182–5. doi: 10.1038/526182a.

10. Miguel E, Camerer C, Casey K, Cohen J, Esterling KM, Gerber A, et al. Social science. Promoting transparency in social science research. Science. 2014;343(6166):30–1. doi: 10.1126/science.1245317.

11. Gorman DM, Ferdinand AO. High impact nutrition and dietetics journals’ use of publication procedures to increase research transparency. Res Integr Peer Rev. 2020;5:12. doi: 10.1186/s41073-020-00098-9.

12. Zuñiga-Hernandez JA, Dorsey-Treviño EG, González-González JG, et al. Endorsement of reporting guidelines and study registration by endocrine and internal medicine journals: meta-epidemiological study. BMJ Open. 2019;9(9):e031259. doi: 10.1136/bmjopen-2019-031259

13. Silva FM, Adegboye ARA, Curioni C, Gomes FS, Collins GS, Kac G, et al. Protocol for a meta-research study of protocols for diet or nutrition-related trials published in indexed journals: general aspects of study design, rationale and reporting limitations. BMJ Open. 2022;12(12):e064744. doi: 10.1136/bmjopen-2022-064744.

14. Naude CE, Durao S, Harper A, Volmink J. Scope and quality of Cochrane reviews of nutrition interventions: a cross-sectional study. Nutr J. 2017;16(1):22. doi: 10.1186/s12937-017-0244-7.

15. Durão S, Kredo T, Volmink J. Validation of a search strategy to identify nutrition trials in PubMed using the relative recall method. J Clin Epidemiol. 2015;68(6):610–6. doi: 10.1016/j.jclinepi.2015.02.005.

16. Madden K, Arseneau E, Evaniew N, Smith CS, Thabane L. Reporting of planned statistical methods in published surgical randomised trial protocols: a protocol for a methodological systematic review. BMJ Open. 2016;6(6):e011188. doi: 10.1136/bmjopen-2016-011188.

17. Ouzzani M, Hammady H, Fedorowicz Z, Elmagarmid A. Rayyan-a web and mobile app for systematic reviews. Syst Rev. 2016;5(1):210. doi: 10.1186/s13643-016-0384-4.

18. RedCAP. Available: https://www.project-redcap.org/ [Accessed 4 Nov 2022].

19. Schulz KF, Altman DG, Moher D; CONSORT Group. CONSORT 2010 statement: updated guidelines for reporting parallel group randomised trials. PLoS Med. 2010;7(3):e1000251. doi: 10.1371/journal.pmed.1000251.

20. Hoffmann TC, Glasziou PP, Boutron I, Milne R, Perera R, Moher D, et al. Better reporting of interventions: template for intervention description and replication (TIDieR) checklist and guide. BMJ. 2014;348:g1687. doi: 10.1136/bmj.g1687

21. Bray F, Laversanne M, Weiderpass E, Soerjomataram I. The ever-increasing importance of cancer as a leading cause of premature death worldwide. Cancer. 2021;127(16):3029–30.

22. Trepanowski JF, Ioannidis JPA. Perspective: Limiting Dependence on Nonrandomized Studies and Improving Randomized Trials in Human Nutrition Research: Why and How. Adv Nutr. 2018;9(4):367–377. doi: 10.1093/advances/nmy014.

23. Schwab S, Janiaud P, Dayan M, Amrhein V, Panczak R, Palagi PM, et al. Ten simple rules for good research practice. PLoS Comput Biol. 2022;18(6):e1010139. doi: 10.1371/journal.pcbi.1010139.

24. Bradley SH, DeVito NJ, Lloyd KE, Richards GC, Rombey T, Wayant C, et al. Reducing bias and improving transparency in medical research: a critical overview of the problems, progress and suggested next steps. J R Soc Med. 2020;113(11):433–443. doi: 10.1177/0141076820956799

25. Macleod MR, Michie S, Roberts I, Dirnagl U, Chalmers I, Ioannidis JPA, et al. Biomedical research: increasing value, reducing waste. Lancet. 2014;383(9912):101–4. doi: 10.1016/S0140-6736(13)62329-6.

26. Soderberg CK, Errington TM, Schiavone SR, Bottesini J, Thorn FS, Vazire S, et al. Initial evidence of research quality of registered reports compared with the standard publishing model. Nat Hum Behav. 202;5(8):990–997. doi: 10.1038/s41562-021-01142-4.

27. Eysenbach G. Peer-review and publication of research protocols and proposals: a role for open access journals. J Med Internet Res. 2004;6(3):e37.

28. Chalmers I, Altman DG. How can medical journals help prevent poor medical research? Some opportunities presented by electronic publishing. Lancet. 1999;353(9151):490–3.

29. Chambers CD, Mellor DT. Protocol transparency is vital for registered reports. Nat Hum Behav. 2018;2(11):791–792. doi: 10.1038/s41562-018-0449-6.

30. Wallach JD, Boyack KW, Ioannidis JPA. Reproducible research practices, transparency, and open access data in the biomedical literature, 2015-2017. PLoS Biol. 2018;16(11):e2006930. doi: 10.1371/journal.pbio.2006930.

31. Serghiou S, Contopoulos-Ioannidis DG, Boyack KW, Riedel N, Wallach JD, Ioannidis JPA. Assessment of transparency indicators across the biomedical literature: How open is open? PLoS Biol. 2021; 19(3): e3001107.

32. Hopewell, S., Boutron, I., Chan, AW. et al. An update to SPIRIT and CONSORT reporting guidelines to enhance transparency in randomized trials. Nat Med. 2022; 28: 1740–1743. doi: 10.1038/s41591-022-01989-8.

33. Schlussel M, Moraes Silva F, the STAR-Nut steering group (2019) Securing Transparency And Reproducibility in studies of NUTritional interventions (STAR-NUT): A research programme to consolidate reporting standards for randomised controlled trials and systematic reviews of nutritional interventions. https://osf.io/b38z9/.

34. Rigutto-Farebrother J, Ahles S, Cade J, Murphy KJ, Plat J, Schwingshackl L, Roche HM, Shyam S, Lachat C, Minihane AM, Weaver C. Perspectives on the application of CONSORT guidelines to randomised controlled trials in nutrition. Eur J Nutr. 2023 Apr 26. doi: 10.1007/s00394-023-03137-5. Epub ahead of print. PMID: 37099211.

