## Supplemental Material for "Describing the landscape of nutrition or diet-related randomised controlled trials: meta-research study of protocols published between 2012 and 2022"

**Authors and affiliations**

Flávia Moraes Silva, Amanda Rodrigues Amorim Adegboye, Cintia Curioni, Fabio Gomes, Gary S Collins, Gilberto Kac, Jonathan Cook, Leila Cheikh Ismail, Matthew J Page, Neha Khandpur, Sarah Sallie Lamb, Sally Hopewell, Shaima Saleh, Shona Kirtley, Simone Bernardes, Solange Durão, Colby J Vorland, Michael Schlussel

#### Supplementary Methods

*Data analysis*

Reporting of funding and conflicts of interest were categorised as: no reporting of funding/ no reporting of conflicts of interest, reporting of no funding/ reporting of no conflicts of interest, and reporting of funding/ reporting of conflicts of interest. We did not investigate the type of funding, the funders' role in the study conduction, nor the type of conflicts of interest, nor verified misreporting.

The clinical conditions of participants were grouped according to the types of diseases, such as endocrine, lung, cardiovascular, gastrointestinal, infectious, muscle and/or skeletal/ rheumatological, psychiatric diseases, kidney, neurological, gynaecological, or others (individual frequency $\leq$ 1.0%).

The intervention category 'complete diet or dietary pattern' details were grouped according to the type of diet, such as Mediterranean Diet, low-carb diet, low-fat diet, healthy diet, personalised diet, intermittent fasting, Dash Diet or sodium-restricted diet, high-quality carb diet, others.

The intervention category 'supplementation, supplements or fortification' was detailed according to the type of supplement as vitamins, minerals, probiotics and synbiotics, carbohydrates and fibre, fats, protein and amino acids, bioactive components, and others.

#### Supplementary Results

*Characteristics of protocols according to the year of publication, countries, cancer and CVD diagnoses*

From 2012 to 2022 there was no discernible trend in the proportion of protocols according to participants' life stages. Adults and the elderly were the most frequent categories in all years except 2012, when adult participants were more common. Individuals with a specific clinical condition were also the most commonly studied groups of individuals, regardless of the year in which the protocol was published. We found no clear relationship between intervention type and the year of protocol publication (**Supplementary Figure 3**). Markers of nutritional status were the most frequent primary outcomes in 2012 and 2013, indicators of frequency or severity of disease in 2015, whereas primary outcomes related to clinical status were more investigated in the remaining years. Regardless of the year of protocol publication, the most frequent study design was the two-arms parallel RCT. The lowest frequencies of protocol registration (85.9%) and conflicts of interest disclosure (81.7%) were observed in 2013, while the frequency of funding disclosure was consistently higher than 95% between 2012 and 2022 (**Supplementary Table 2).**

**Supplementary Table 3** shows the frequency of each component of the PICOS acronym for nutrition or diet-related RCTs' protocols and practices of transparency and reproducibility in the five most frequent countries where the study would be conducted (the USA, UK, Iran, China, and Australia). Interventions related to nutrition education were more common in the USA (50.9%), whereas interventions related to supplementation were more frequent in other countries, ranging from 38.5% in China to 75.5% in Iran. It was also the most frequent intervention in Canada, Germany and Netherlands while in Brazil it was similar to interventions related to nutrition education as illustrated in **Supplementary Figure 4.**

The type of comparator was heterogeneous among these countries: other interventions were more frequent in the USA (38.2%), usual care was more common in Australia (36.5%), while placebo was the most frequent comparator in the other countries, and it ranged from 35.4% (China) to 73.8% (Iran). Adults and the elderly (23.0% [USA] to 40.3% [UK]) with a specific clinical condition (61.8% [USA] to 89.2% [Iran]) were more commonly included in the protocols, regardless of the country. The most frequent primary outcome was related to nutritional status in Chinese protocols (37.6%), withdrawal or adherence in UK protocols (22.2%), while in the other countries, it was mainly related to clinical status (24.1% [Australia] to 75.4% [Iran]). Protocols in the USA were less likely to report registration information (83%) and a conflict of interest statement (66.4%), whereas these practices of transparency were more frequent in protocols of trials conducted in Iran (98.5% for both). SPIRIT (55.4%), TIDiER (8.3%), and CONSORT (40.9%) were more mentioned by protocols from Iran, the UK, and Australia, respectively.

The characteristics of the protocols involving patients with cancer and CVD disease are presented in **Supplementary Table 4**. Both subgroups involved mainly adults and the elderly as participants and most frequently had an intervention related to supplementation. Primary outcomes related to the clinical status were more frequent in protocols involving patients with CVD diseases (43.3%), while the incidence or severity of disease was more common in protocols involving patients with cancer (25.0%). Unfunded RCTs comprised 10.5% and 1.1% of protocols involving patients with cancer and CVD disease, respectively, whereas the frequency of protocols reporting conflicts of interest was similar (~14%) in both subgroups. Only 2.2% of protocols involving patients with CVD diseases did not report to the protocol registry, while 9.2% of protocols involving patients with cancer did not do this.

#### Data extraction form

### Draft Data Extraction Protocols Of Nutrition or Diet-related RCTs

| | |
|---|---|
| Number of report | __________________________ |
| Confirmation of eligibility | ◯ no<br>◯ yes |
| Reason of no eligibility: | ◯ it is not a protocol<br>◯ intervention is not related to nutrition or diet<br>◯ it is not a protocol of RCT<br>◯ it is a preprint<br>◯ it is an abstract<br>◯ it is a duplicate |
| First author | __________________________ |
| Year of publication | __________________________ |
| Journal of publication | __________________________ |
| PICOTS_ population | ☐ Pregnant women<br>☐ Mother and infant pairs<br>☐ Infants<br>☐ Children and preschool-aged children<br>☐ Adolescents<br>☐ Adults<br>☐ Elderly<br>☐ Adults and elderly<br>☐ Postmenopausal women<br>☐ Participants with a clinical condition (s)<br>☐ Families |
| PICOTS_ population_clinical condition | __________________________ |
| PICOTS_ population_cancer | ◯ no<br>◯ yes |
| PICOTS_ population_CVD | ◯ no<br>◯ yes |

| | |
|---|---|
| PICOTS_intervention_complexity | ◯ Only nutrition or diet-related intervention<br>◯ Nutrition or diet-related intervention combined with exercise<br>◯ Nutrition or diet-related intervention combined with drugs<br>◯ Nutrition or diet-related intervention combined with other medical care<br>◯ Nutrition or diet-related intervention as part of a lifestyle intervention<br>◯ Nutrition or diet-related intervention as part of a health intervention |
| PICOTS_intervention_type | ◯ Food (whole food, food products, specially formulated foods)<br>◯ Breastfeeding, complementary feeding, weaning<br>◯ Complete diet or dietary patterns<br>◯ Complete nutrition formulas (enteral or parenteral)<br>◯ Supplementation or supplements (single or multiple nutrients, bioactive non-nutrients, plant components)<br>◯ Nutrition education, counseling and coordination of care<br>◯ Other, if no component of intervention could be categorized as any of the above |
| PICOTS_intervention_type_details | __________________________________ |
| Number of arms (groups) | ◯ 2<br>◯ 3<br>◯ 4<br>◯ 5 or more |
| PICOTS_Control | ◯ Placebo<br>◯ Usual care<br>◯ Other intervention<br>◯ No control (such as wait list) |
| PICOTS_duration | __________________________________<br>(in months) |
| PICOTS_primary outcome | ◯ not specified<br>◯ specified |

| | |
|---|---|
| primary outcome | ☐ Mortality<br>☐ Clinical status (clinical or biochemical measures)<br>☐ Nutritional status (anthropometry, body composition, nutrition diagnosis)<br>☐ Functional status<br>☐ Frequency or severity of disease<br>☐ Diet quality and/or variety<br>☐ Food/ nutrient/ dietary intake<br>☐ Diet-related behaviors<br>☐ Other non-dietary behaviors<br>☐ Withdrawal from the study, drop-out or adherence-related<br>☐ Adverse events, side-effects and/or safety<br>☐ Cost-effectiveness or economic<br>☐ Quality of life<br>☐ Breastfeeding<br>☐ Other |
| other primary outcome description | __________________________________ |
| Study Design | ○ Parallel<br>○ Crossover |
| Pilot study | ○ No<br>○ Yes |
| Study Design_1 | ○ Cluster RCT<br>○ No cluster RCT |
| Study Design_2 | ○ Factorial RCT<br>○ No factorial RCT |
| Study Design_Framework | ○ Superiority<br>○ Non - inferiority<br>○ Equivalence<br>○ Exploratory<br>○ Not reported |
| Study local | ○ unicentric<br>○ multicentric |
| Study country | __________________________________ |
| Registered protocol | ○ no<br>○ yes |
| Reference to SPIRIT | ○ no<br>○ yes |
| Reference to CONSORT | ○ no<br>○ yes |
| Reference to TIDieR | ○ no<br>○ yes |





| | |
|---|---|
| Reference to CONSERVE | ○ no<br>○ yes |

**Supplementary Box 1**. Search Strategy

**PubMed**

Database and platform: PubMed 1946 to present (via https://pubmed.ncbi.nlm.nih.gov/)

Search filter: Cochrane Highly Sensitive Search Strategy for identifying randomised trials in MEDLINE: sensitivity- and precision-maximizing version (2008 revision); PubMed format (https://handbook-5-1.cochrane.org/chapter_6/box_6_4_b_cochrane_hsss_2008_sensprec_pubmed.htm)

#1
("Randomized controlled trial" [pt] OR "controlled clinical trial" [pt] OR randomized [tiab] OR randomised [tiab] OR placebo [tiab] OR clinical trial as topic [mesh:noexp] OR randomly [tiab] OR trial [ti]) NOT (animals [mh] NOT humans [mh])
#2
(Nutritional Sciences[mh] OR Nutritional Physiological Phenomena[mh] OR Nutrition Assessment[mh] OR Nutrition Therapy[mh] OR Nutritional and Metabolic Diseases[mh] OR nutrition*[tiab] OR diet[tiab] OR feeding[tiab] OR dietary[tiab] OR breastfeed*[tiab] OR breast feed*[tiab] OR lactation[tiab] OR bottle feed*[tiab] OR complementary feeding[tiab] OR weaning[tiab] OR enteral[tiab] OR parenteral[tiab] OR Feeding Methods[mh] OR nutritional status[tiab] OR overweight[tiab] OR obese[tiab] OR obesity[tiab] OR overnutrition[tiab] OR over nutrition[tiab] OR undernourished[tiab] OR overnourished[tiab] OR wasted[tiab] OR wasting[tiab] OR stunting[tiab] OR stunted[tiab] OR underweight[tiab] OR undernutrition[tiab] OR under nutrition[tiab] OR body weight[tiab] OR anthropometry[tiab] OR Body Weights and Measures[mh] OR growth monitoring[tiab] OR food[tiab] OR food labelling[mh] OR food assistance[mh] OR supplementary feeding[tiab] OR diet therapy[mh] OR food and beverages[mh] OR vegetable*[tiab] OR fruit*[tiab] OR meat[tiab] OR dairy[tiab] OR dietary fat*[tiab] OR starch*[tiab] OR cereal[tiab] OR food-drug interactions[mh] OR food supply[mh] OR feeding behavio*[tiab] OR eating behavio*[tiab] OR food pattern*[tiab] OR food hypersensitivity[mh] OR food deprivation[mh] OR food, organic [mh] OR micronutrient*[tiab] OR vitamin*[tiab] OR thiamin[tiab] OR riboflavin[tiab] OR niacin[tiab] OR pantothenic acid[tiab] OR pyridoxine[tiab] OR pyridoxal[tiab] OR pyridoxamine[tiab] OR biotin[tiab] OR folic acid[tiab] OR folate[tiab]OR cyanocobalamin[tiab] OR choline[tiab] OR retinol[tiab] OR ascorbic acid[tiab] OR tocopherol[tiab] OR carotenoids[tiab] OR carotene[tiab] OR cryptoxanthin[tiab] OR lutein[tiab] OR lycopene[tiab] OR zeaxanthin[tiab] OR minerals[tiab] OR calcium[tiab] OR chloride[tiab] OR magnesium[tiab] OR phosphorus[tiab] OR potassium[tiab] OR sodium[tiab] OR sulphur[tiab] OR trace element*[tiab] OR boron[tiab] OR cobalt[tiab] OR chromium[tiab] OR copper[tiab] OR fluoride[tiab] OR iodine[tiab] OR iron[tiab] OR manganese[tiab] OR molybdenum[tiab] OR selenium[tiab] OR zinc[tiab] OR trace metal*[tiab] OR macronutrient*[tiab] OR carbohydrate*[tiab] OR dietary protein*[tiab] OR saturated fat*[tiab] OR unsaturated fat*[tiab] OR mono unsaturated fat*[tiab] OR monounsaturated fat*[tiab] OR poly unsaturated fat*[tiab] OR polyunsaturated fat*[tiab] OR trans fat*[tiab] OR dietary fibre[tiab] OR dietary fiber[tiab] OR dietary salt[tiab] OR table salt[tiab] OR soft drink[tiab] OR fruit juice[tiab] OR vegetable juice[tiab] OR milk[tiab] OR tea[tiab] OR coffee[tiab] OR energy drink*[tiab] OR carbonated beverage*[tiab] OR carbonated drink*[tiab] OR prebiotics[tiab] OR probiotics[tiab] OR glycemic load[tiab] OR glycemic index[tiab] OR calories[tiab] OR kilocalories[tiab] OR kilojoules[tiab] OR caloric intake[tiab] OR energy intake[tiab]

#3
(protocol*[ti] OR study design [ti] OR trial design[ti] OR research design[ti] OR "design and methods"[ti] OR "design and rationale"[ti] OR "rationale and design"[ti] OR Research Design[mh] OR clinical protocols[mh] OR clinical trial protocol[pt])
#4
#1 AND #2 AND #3
#4 *Filters applied: From 2012/1/1 to 2022/3/24.*

**Embase**

Database and platform: Embase 1947 to present (via Elsevier)
Search filter: SIGN RCT filter
https://www.sign.ac.uk/what-we-do/methodology/search-filters/

#7
#6 AND (2012:py OR 2013:py OR 2014:py OR 2015:py OR 2016:py OR 2017:py OR 2018:py OR 2019:py OR 2020:py OR 2021:py OR 2022:py)
#6
#1 AND #4 AND #5
#5
"Methodology"/de OR "Clinical Protocol"/exp OR protocol:ti OR 'study design':ti OR "design and methods":ti OR "design and rationale":ti OR "rationale and design":ti
#4
#2 NOT #3
#3
'case study'/de OR 'case report':ab,ti OR 'abstract report'/de OR letter/de OR 'conference paper':it OR 'conference abstract':it OR editorial:it OR letter:it OR note:it
#2
'clinical trial'/de OR 'randomized controlled trial'/de OR 'controlled clinical trial'/de OR 'multicenter study'/de OR 'phase 3 clinical trial'/de OR 'phase 4 clinical trial'/de OR randomization/exp OR 'single blind procedure'/de OR 'double blind procedure'/de OR 'crossover procedure'/de OR placebo/de OR 'randomi?ed controlled trial*$':ab,ti OR rct:ab,ti OR 'random* NEAR/2 allocat*':ab,ti OR 'single blind*':ab,ti OR 'double blind*':ab,ti OR ((treble:ab,ti OR triple:ab,ti) NEAR blind*:ab,ti) OR placebo*:ab,ti OR 'prospective study'/de
#1
'Nutritional Science'/exp OR 'nutritional science':kw OR 'nutritional physiological phenomena':ab,ti OR 'Nutrition'/exp OR 'nutritional assessment':kw OR 'nutritional support':kw OR 'diet therapy':kw OR 'Diet Therapy'/exp OR (nutritional:kw AND 'metabolic disorder':kw) OR 'Nutritional Disorder'/exp OR 'Metabolic Disorder'/exp OR nutrition:ab,ti OR diet:ab,ti,kw OR feeding:ab,ti OR 'Food'/exp OR 'dietary intake':ab,ti OR 'Diet Restriction'/exp OR breastfeeding:ab,ti OR lactation:ab,ti OR 'bottle feeding':ab,ti OR 'complementary feeding':ab,ti OR weaning:ab,ti OR 'enteric feeding':ab,ti OR parenteral:ab,ti OR 'food intake':ab,ti OR 'nutritional status':ab,ti OR 'Failure to Thrive'/de OR 'Body Weight'/exp OR obesity:ab,ti OR 'obese patient':ab,ti OR overnutrition:ab,ti OR 'wasting syndrome':ab,ti OR stunting:ab,ti OR 'stunting syndrome':ab,ti OR malnutrition:ab,ti OR 'malnutrition'/de OR 'body weight':ab,ti OR anthropometry:kw OR ('growth, development':ab,ti AND aging:ab,ti) OR food:ab,ti OR 'food packaging':kw OR 'Food Packaging'/de OR 'food assistance':kw OR 'Food Assistance'/de OR 'supplementary feeding':ab,ti OR 'Food Insecurity'/exp OR vegetable:ab,ti OR fruit:ab,ti OR meat:ab,ti OR dairy:ab,ti OR 'fat intake':ab,ti OR starch:ab,ti OR cereal:ab,ti OR 'food drug interaction':ab,ti OR 'catering service':kw OR 'feeding

behavior':ab,ti OR 'eating habit':ab,ti OR 'food allergy':kw OR 'food deprivation':kw OR ‘Food Deprivation’/exp OR 'organic food':ab,ti OR ‘Organic Food’/de OR 'micronutrient intake':ab,ti OR vitamin:ab,ti OR thiamine:ab,ti OR riboflavin:ab,ti OR 'nicotinic acid':ab,ti OR 'pantothenic acid':ab,ti OR pyridoxine:ab,ti OR pyridoxal:ab,ti OR pyridoxamine:ab,ti OR biotin:ab,ti OR 'folic acid':ab,ti OR cyanocobalamin:ab,ti OR choline:ab,ti OR retinol:ab,ti OR 'ascorbic acid':ab,ti OR tocopherol:ab,ti OR carotenoid:ab,ti OR carotene:ab,ti OR cryptoxanthin:ab,ti OR xanthophyll:ab,ti OR lycopene:ab,ti OR zeaxanthin:ab,ti OR mineral:ab,ti OR calcium:ab,ti OR chloride:ab,ti OR magnesium:ab,ti OR phosphorus:ab,ti OR potassium:ab,ti OR sodium:ab,ti OR sulfur:ab,ti OR 'trace element':ab,ti OR boron:ab,ti OR cobalt:ab,ti OR chromium:ab,ti OR copper:ab,ti OR fluoride:ab,ti OR iodine:ab,ti OR iron:ab,ti OR manganese:ab,ti OR molybdenum:ab,ti OR selenium:ab,ti OR zinc:ab,ti OR 'trace metal':ab,ti OR macronutrient:ab,ti OR carbohydrate:ab,ti OR 'protein intake':ab,ti OR 'saturated fatty acid':ab,ti OR 'unsaturated fatty acid':ab,ti OR 'monounsaturated fatty acid':ab,ti OR 'polyunsaturated fatty acid':ab,ti OR 'trans fatty acid':ab,ti OR 'dietary fiber':ab,ti OR 'salt intake':ab,ti OR 'soft drink':ab,ti OR 'fruit juice':ab,ti OR 'vegetable juice':ab,ti OR milk:ab,ti OR tea:ab,ti OR coffee:ab,ti OR 'energy drink':ab,ti OR 'carbonated beverage':ab,ti OR 'prebiotic agent':ab,ti OR 'probiotic agent':ab,ti OR 'glycemic index':ab,ti OR 'glycemic load':ab,ti OR calorie:ab,ti OR 'caloric intake':ab,ti

**Web of Science**

Database and platform: Web of Science (All databases) 1945 to present (via Clarivate)
Search filter: Cochrane ENT group RCT filter
https://ent.cochrane.org/sites/ent.cochrane.org/files/public/uploads/rct_filters.pdf

#1
(((((((TS=(protocol*)) OR TS=("study design")) OR TS=("trial design")) OR TS=("research design")) OR TS=("design and methods")) OR TS=("design and rationale")) OR TS=("rationale and design"))
#2
TS=(randomised OR randomized OR randomisation OR randomisation OR placebo* OR (random*))
#3
(((((((((((((((((((((((((((((((((((((((((((((((((((((((((((((((((((((((((((((((((((((((((((((((((((((((((((((((((((((((((((((((((((((((((((((((((((((((((((((( ((((((((((((((AK=("nutritional sciences")) OR AK=("nutritional physiological phenomena" )) OR AK=("nutrition assessment" )) OR AK=("nutritional support")) OR AK=(“nutrition”)) OR AK=("nutrition therapy")) OR AK=( "nutritional and metabolic diseases" )) OR TS=( "nutrition*")) OR KP=(“nutrition”)) OR KP=(“diet”)) OR TS =("diet" )) OR TS =("feeding" )) OR TS =("dietary" )) OR TS =( "breastfeed*" )) OR TS =( "breast feed*")) OR TS =( "lactation" )) OR TS =( "bottle feed*" )) OR TS =("complementary feeding")) OR TS =("weaning" )) OR TS =("enteral" )) OR TS =( "parenteral")) OR AK=("feeding methods" )) OR TS =( "nutritional status" )) OR KP=(“nutrition support”)) OR TS =("overweight" )) OR TS =("obese" )) OR TS =("obesity" )) OR TS =("overnutrition")) OR TS =( "over nutrition")) OR TS =("undernourished")) OR TS =("overnourished")) OR TS =("wasted")) OR TS =( "wasting")) OR TS =("stunting")) OR TS =( "stunted")) OR TS =( "underweight")) OR TS =( "undernutrition")) OR TS =("under nutrition")) OR TS =("body weight")) OR TS =( "anthropometry")) OR AK=( "body weights and measures")) OR TS =( "growth monitoring")) OR TS =("food")) OR AK=( "food labeling")) OR AK=( "food assistance")) OR TS =("supplementary feeding")) OR AK=( "diet therapy")) OR AK=("food and beverages")) OR TS =( "vegetable*")) OR TS =("fruit*")) OR TS =( "meat")) OR TS =("dairy")) OR TS =( "dietary fat*")) OR AB=("starch*")) OR

TS =( "cereal")) OR AK=( "food drug interactions")) OR AK=("food supply")) OR TS =("feeding behavio*")) OR TS =( "eating behavio*")) OR TS =( "food pattern*")) OR TS =("food hypersensitivity")) OR TS =( "food deprivation")) OR TS =( "food, organic")) OR TS =( "micronutrient*")) OR TS =("vitamin*")) OR TS =( "thiamin")) OR TS =( "riboflavin")) OR TS =( "niacin")) OR TS =( "pantothenic acid")) OR TS =("pyridoxine")) OR TS =( "pyridoxal")) OR TS =( "pyridoxamine")) OR TS =( "biotin")) OR TS =( "folic acid")) OR TS =("folate")) OR TS =("cyanocobalamin")) OR TS =("choline")) OR TS =("retinol")) OR TS =("ascorbic acid")) OR TS =("tocopherol")) OR TS =("carotenoids")) OR TS =("carotene")) OR TS =("cryptoxanthin")) OR TS =("lutein")) OR TS =("lycopene")) OR TS =("zeaxanthin")) OR TS =("minerals")) OR TS =("calcium")) OR TS =("chloride")) OR TS =("magnesium")) OR TS =("phosphorus")) OR TS =("potassium")) OR TS =("sodium")) OR TS =("sulphur")) OR TS =("trace element*")) OR TS =("boron")) OR TS =("cobalt")) OR TS =("chromium")) OR TS =("copper")) OR TS =("fluoride")) OR TS =("iodine")) OR TS =("iron")) OR TS =("manganese")) OR TS =("molybdenum")) OR TS =("selenium")) OR TS =("zinc")) OR TS =("trace metal*")) OR TS =("macronutrient*")) OR TS =("carbohydrate*")) OR TS =("dietary protein*")) OR TS =("saturated fat*")) OR TS =("unsaturated fat*" )) OR TS =("mono unsaturated fat*")) OR TS =("monounsaturated fat*")) OR TS =("poly unsaturated fat*")) OR TS =("polyunsaturated fat*")) OR TS =("trans fat*")) OR TS =("dietary fibre")) OR TS =("dietary fiber")) OR TS =("dietary salt")) OR TS =("table salt")) OR TS =("soft drink")) OR TS =("fruit juice")) OR TS =("vegetable juice")) OR TS =("milk")) OR TS =("tea")) OR TS =("coffee")) OR TS =("energy drink*")) OR TS =("carbonated beverage*")) OR TS =("carbonated drink*")) OR TS =("prebiotics")) OR TS =("probiotics")) OR TS =("glycaemic load")) OR TS =("glycaemic index")) OR TS =("calories")) OR TS =("kilocalories")) OR TS =("kilojoules")) OR TS =("caloric intake")) OR TS =("energy intake")

#4

#1 AND #2 AND #3

#5

#4 Timespan: 2012-01-01 to 2022-03-24 (Publication Date)

**CINAHL**

Database and platform: CINAHL with full text 1981 to present (via EBSCOHost)

Search filter: SIGN CINAHL for EBSCO RCT Filter (created by Mark Clowes)
https://www.sign.ac.uk/assets/search-filters-randomised-controlled-trials.docx

S1

((MH "Research Protocols") OR (MH "Research Methodology") OR (MH "Study Design") ) OR PT Protocol OR ( TI (protocol* OR "study design" OR "trial design" OR "research design" OR "design and methods" OR "design and rationale" OR "rationale and design") )

S2

((MH "Nutrition+") OR (MH "Nutritional Physiology+") OR (MH "Nutritional Assessment") OR (MH "Diet Therapy+") OR (MH "Nutritional and Metabolic Diseases+") OR (MH "Feeding Methods") OR (MH “Diet+”) ) OR ( TI (nutrition* OR diet OR feeding OR dietary OR breastfeed* OR "breast feed*" OR lactation OR "bottle feed*" OR "complementary feeding" OR weaning OR enteral OR parenteral) OR AB (nutrition* OR diet OR feeding OR dietary OR breastfeed* OR "breast feed*" OR lactation OR "bottle feed*" OR "complementary feeding" OR weaning OR enteral OR parenteral) ) OR ( TI ("nutritional status" OR overweight OR obese OR obesity OR overnutrition OR "over nutrition" OR undernourished OR overnourished OR wasted OR wasting OR stunting OR stunted OR underweight

OR undernutrition OR "under nutrition" OR "body weight" OR anthropometry) OR AB ("nutritional status" OR overweight OR obese OR obesity OR overnutrition OR "over nutrition" OR undernourished OR overnourished OR wasted OR wasting OR stunting OR stunted OR underweight OR undernutrition OR "under nutrition" OR "body weight" OR anthropometry) ) OR ( (MH "Body Weights and Measures") OR (MH "Anthropometry") OR (MH "Body Weight") OR (MH "Food Labeling") OR (MH "Food Assistance") OR (MH "Food and Beverages+") ) OR ( TI ("growth monitoring" OR food OR "supplementary feeding" OR vegetable* OR fruit* OR meat OR dairy OR "dietary fat*" OR starch* OR cereal) OR AB ("growth monitoring" OR food OR "supplementary feeding" OR vegetable* OR fruit* OR meat OR dairy OR "dietary fat*" OR starch* OR cereal) ) OR ( (MH "Drug-Food Interactions") OR (MH "Food Supply") OR (MH "Organic Food") OR (MH "Food Hypersensitivity") ) OR ( TI ("feeding behavio*" OR "eating behavio*" OR "food pattern*" OR micronutrient* OR vitamin* OR thiamin OR riboflavin OR niacin OR "pantothenic acid" OR pyridoxine OR pyridoxal OR pyridoxamine OR biotin OR "folic acid" OR folate OR cyanocobalamin OR choline OR retinol OR "ascorbic acid" OR tocopherol OR carotenoids OR carotene OR cryptoxanthin) OR AB ("feeding behavio*" OR "eating behavio*" OR "food pattern*" OR micronutrient* OR vitamin* OR thiamin OR riboflavin OR niacin OR "pantothenic acid" OR pyridoxine OR pyridoxal OR pyridoxamine OR biotin OR "folic acid" OR folate OR cyanocobalamin OR choline OR retinol OR "ascorbic acid" OR tocopherol OR carotenoids OR carotene OR cryptoxanthin) ) OR ( TI (lutein OR lycopene OR zeaxanthin OR minerals OR calcium OR chloride OR magnesium OR phosphorus OR potassium OR sodium OR sulphur OR "trace element*" OR boron OR cobalt OR chromium OR copper OR fluoride OR iodine OR iron OR manganese OR molybdenum OR selenium OR zinc OR "trace metal*" OR macronutrient* OR carbohydrate* OR "dietary protein*") OR AB (lutein OR lycopene OR zeaxanthin OR minerals OR calcium OR chloride OR magnesium OR phosphorus OR potassium OR sodium OR sulphur OR "trace element*" OR boron OR cobalt OR chromium OR copper OR fluoride OR iodine OR iron OR manganese OR molybdenum OR selenium OR zinc OR "trace metal*" OR macronutrient* OR carbohydrate* OR "dietary protein*") ) OR ( TI ("saturated fat*" OR "unsaturated fat*" OR "mono unsaturated fat*" OR "monounsaturated fat*" OR "poly unsaturated fat*" OR "polyunsaturated fat*" OR "trans fat*" OR "dietary fibre" OR "dietary fiber" OR "dietary salt" OR "table salt" OR "soft drink" OR "fruit juice" OR "vegetable juice") OR AB ("saturated fat*" OR "unsaturated fat*" OR "mono unsaturated fat*" OR "monounsaturated fat*" OR "poly unsaturated fat*" OR "polyunsaturated fat*" OR "trans fat*" OR "dietary fibre" OR "dietary fiber" OR "dietary salt" OR "table salt" OR "soft drink" OR "fruit juice" OR "vegetable juice") ) OR ( TI (milk OR tea OR coffee OR "energy drink*" OR "carbonated beverage*" OR "carbonated drink*" OR prebiotics OR probiotics OR "glycemic load" OR "glycemic index" OR calories OR kilocalories OR kilojoules OR "caloric intake" OR "energy intake") OR AB (milk OR tea OR coffee OR "energy drink*" OR "carbonated beverage*" OR "carbonated drink*" OR prebiotics OR probiotics OR "glycemic load" OR "glycemic index" OR calories OR kilocalories OR kilojoules OR "caloric intake" OR "energy intake") )

S3

TX allocat* random* OR (MH "Quantitative Studies") OR (MH "Placebos") OR TX placebo* OR TX random* allocat* OR (MH "Random Assignment") OR TX randomi* control* trial* OR ( TX ( (singl* n1 blind*) or (singl* n1 mask*) ) or TX ( (doubl* n1 blind*) or (doubl* n1 mask*) ) or TX ( (tripl* n1 blind*) or (tripl* n1 mask*) ) or TX ( (trebl* n1 blind*) or (trebl* n1 mask*) ) ) OR TX clinic* n1 trial* OR PT Clinical trial OR (MH "Clinical Trials+")

S4

S1 AND S2 AND S3

S5

S4 (LIMITADORES 202203 - 201201)

**Global Health**

Database and platform: Global Health 1973 to 2022 Week 12 (via OVID)

Search filter: RCT filter adapted from the Cochrane Highly Sensitive Search Strategy for identifying randomised trials in MEDLINE: sensitivity- and precision-maximising version (2008 revision)

1. (protocol$ OR "study design$" OR "trial design$" OR "research design$" OR "design and methods" OR "design and rationale" OR "rationale and design").ti.

2. Methodology/ OR Experimental Design/

3. 1 OR 2

4. exp Nutrition/ OR exp Nutrition Research/ OR exp Human Feeding/ OR Diets/ OR Diet/

5. Nutrition Physiology/ OR Nutritional Assessment/ OR Nutritional Disorders/ OR Nutritional Intervention/

6. (nutrition$ OR diet OR feeding OR dietary OR breastfeed$ OR "breast feed$" OR lactation OR "bottle feed$" OR "complementary feeding" OR weaning OR enteral OR parenteral).ti,ab.

7. ("nutritional status" OR overweight OR obese OR obesity OR overnutrition OR "over nutrition" OR undernourished OR overnourished OR wasted OR wasting OR stunting OR stunted OR underweight OR undernutrition OR "under nutrition" OR "body weight" OR anthropometry).ti,ab.

8. Body Weight/ OR Nutrition Labelling/ OR Diet Treatment/ OR Food Supply/ OR Nutrient Drug Interactions/ OR Food Deprivation/

9. exp Body Measurements/ OR exp Foods/ OR exp Therapeutic Diets/ OR exp Food Allergies/

10. ("growth monitoring" OR food OR "food assistance" OR "supplementary feeding" OR vegetable$ OR fruit$ OR meat$ OR dairy$ OR "dietary fat$" OR starch$ OR cereal OR "feeding behavio$" OR "eating behavio$" OR "food pattern$").ti,ab.

11. ("food hypersensitivit$" OR "organic food" OR micronutrient$ OR vitamin$ OR thiamin OR riboflavin OR niacin OR "pantothenic acid" OR pyridoxine OR pyridoxal OR pyridoxamine OR biotin OR "folic acid" OR folate OR cyanocobalamin OR choline OR retinol OR "ascorbic acid" OR tocopherol OR carotenoids OR carotene OR cryptoxanthin OR lutein OR lycopene OR zeaxanthin OR minerals OR calcium OR chloride OR magnesium OR phosphorus OR potassium OR sodium OR sulphur OR "trace element$" OR boron OR cobalt OR chromium OR copper OR fluoride OR iodine OR iron OR manganese OR molybdenum OR selenium OR zinc OR "trace metal$").ti,ab.

12. (macronutrient$ OR carbohydrate$ OR "dietary protein$" OR "saturated fat$" OR "unsaturated fat$" OR "mono unsaturated fat$" OR "monounsaturated fat$" OR "poly unsaturated fat$" OR "polyunsaturated fat$" OR "trans fat$" OR "dietary fibre" OR "dietary fiber" OR "dietary salt" OR "table salt" OR "soft drink" OR "fruit juice" OR "vegetable juice" OR milk OR tea OR coffee OR "energy drink$" OR "carbonated beverage$" OR "carbonated drink$" OR prebiotics OR probiotics

OR "glycemic load" OR "glycemic index" OR calories OR kilocalories OR kilojoules OR "caloric intake" OR "energy intake").ti,ab.

13. 4 OR 5 OR 6 OR 7 OR 8 OR 9 OR 10 OR 11 OR 12

14. Randomized controlled trials/ OR Clinical Trials/ OR Placebos/

15. (randomized OR randomised OR randomly OR placebo OR trial$).ti,ab.

16. 14 OR 15

17. 3 AND 13 AND 16

18. Limit 17 to yr="2012-2022"

**PsycInfo**

Database and platform: PsycINFO 1806 to present (via OVID)

Search filter: Cochrane ENT Group RCT search filter for PsycInfo (OVID) https://ent.cochrane.org/sites/ent.cochrane.org/files/public/uploads/rct_filters.pdf

1. (protocol$ OR "study design$" OR "trial design$" OR "research design$" OR "design and methods" OR "design and rationale" OR "rationale and design").ti.

2. ("Research Design" OR "clinical protocols").mh.

3. Methodology/

4. Experimental Design/

5. 1 OR 2 OR 3 OR 4

6. ("Nutritional Sciences" OR "Nutritional Physiological Phenomena" OR "Nutrition Assessment" OR "Nutrition Therapy" OR "Nutritional and Metabolic Diseases" OR "Feeding Methods" OR "Body Weights and Measures" OR "Food Labelling" OR "Food Assistance" OR "Diet Therapy" OR "Food and Beverages" OR "Food-drug Interactions" OR "Food Supply" OR "Food Hypersensitivity" OR "Food Deprivation" OR "Food, Organic").mh.

7. exp Nutrition/ OR exp "Nutritional Deficiencies"/ OR exp "Metabolism Disorders"/ OR exp Diets/ OR exp Food/

8. "Eating Behavior"/ OR "Failure to Thrive"/ OR "Body Weight"/ OR "Beverages (Nonalcoholic)"/ OR "Food Insecurity"/ OR "Food Allergies"/ OR "Food Deprivation"/ OR "Food Intake"/ OR "Dietary Supplements"/ OR "Dietary Restraint"/ OR "Food Preferences"/

9. (nutrition$ OR diet OR feeding OR dietary OR breastfeed$ OR "breast feed$" OR lactation OR "bottle feed$" OR "complementary feeding" OR weaning OR enteral OR parenteral OR "nutritional status" OR overweight OR obese OR obesity OR overnutrition OR "over nutrition" OR undernourished OR overnourished OR wasted OR wasting OR stunting OR stunted OR underweight OR undernutrition OR "under nutrition" OR "body weight" OR anthropometry).ti,ab.

10. ("growth monitoring" OR food OR "supplementary feeding" OR vegetable$ OR

fruit$ OR meat OR dairy OR "dietary fat$" OR starch$ OR cereal OR "feeding behavio$" OR "eating behavio$" OR "food pattern$" OR micronutrient$).ti,ab.

11. (vitamin$ OR thiamin OR riboflavin OR niacin OR pantothenic acid OR pyridoxine OR pyridoxal OR pyridoxamine OR biotin OR folic acid OR folate OR cyanocobalamin OR choline OR retinol OR ascorbic acid OR tocopherol OR carotenoids OR carotene OR cryptoxanthin OR lutein OR lycopene OR zeaxanthin OR minerals OR calcium OR chloride OR magnesium OR phosphorus OR potassium OR sodium OR sulphur OR "trace element$" OR boron OR cobalt OR chromium OR copper OR fluoride OR iodine OR iron OR manganese OR molybdenum OR selenium OR zinc OR "trace metal$").ti,ab.

12. (macronutrient$ OR carbohydrate$ OR "dietary protein$" OR "saturated fat$" OR "unsaturated fat$" OR "mono unsaturated fat$" OR "monounsaturated fat$" OR "poly unsaturated fat$" OR "polyunsaturated fat$" OR "trans fat$" OR "dietary fibre" OR "dietary fiber" OR "dietary salt" OR "table salt").ti,ab.

13. ("soft drink" OR "fruit juice" OR "vegetable juice" OR milk OR tea OR coffee OR "energy drink$" OR "carbonated beverage$" OR "carbonated drink$" OR prebiotics OR probiotics OR "glycemic load" OR "glycemic index" OR calories OR kilocalories OR kilojoules OR "caloric intake" OR "energy intake").ti,ab.

14. 6 OR 7 OR 8 OR 9 OR 10 OR 11 OR 12 OR 13

15. Placebo/

16. ((blind* or mask*) and (single or double or triple or treble)).tw.

17. (randomised or randomized or randomisation or randomization or (random* and (allocat* or assign*)) or factorial* or placebo* or crossover* or (cross adj over*)).tw.

18. 15 or 16 or 17

19. 5 AND 14 AND 18

20. limit 19 to yr="2012-2022"

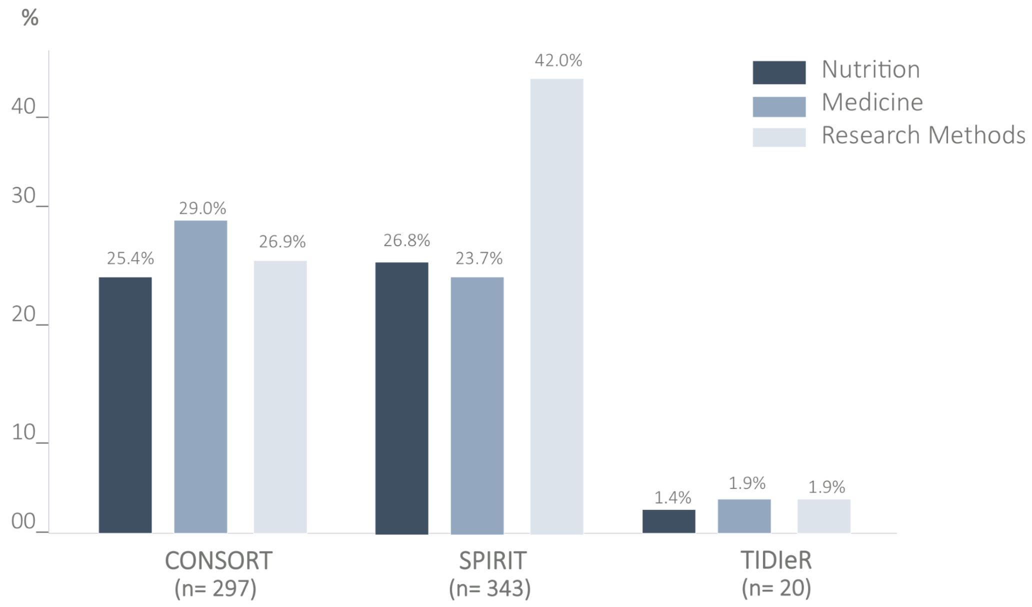


**Supplementary Figure 1**. Relative frequency of published nutrition or diet related RCT protocols that referenced the reporting guidelines according to the scientific field of journals.

**2A**

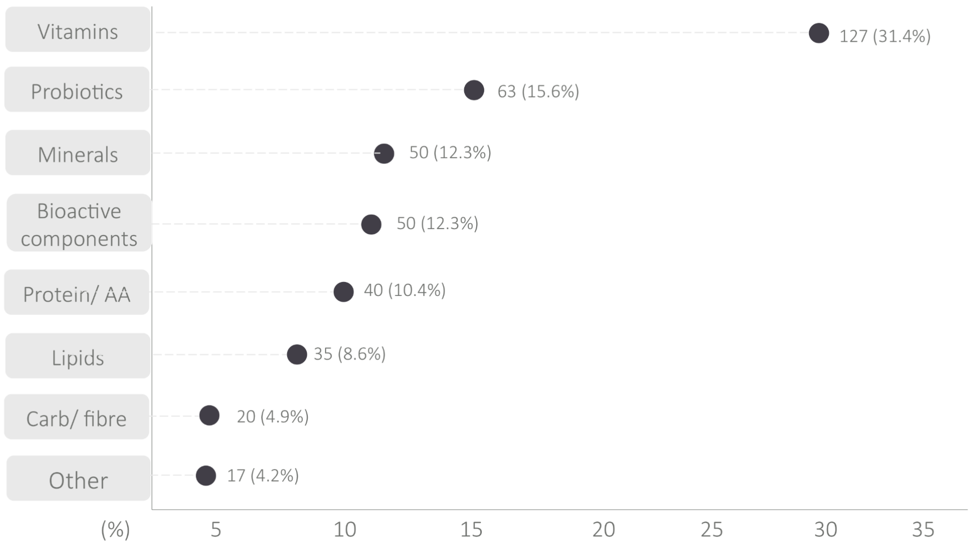


**2B**

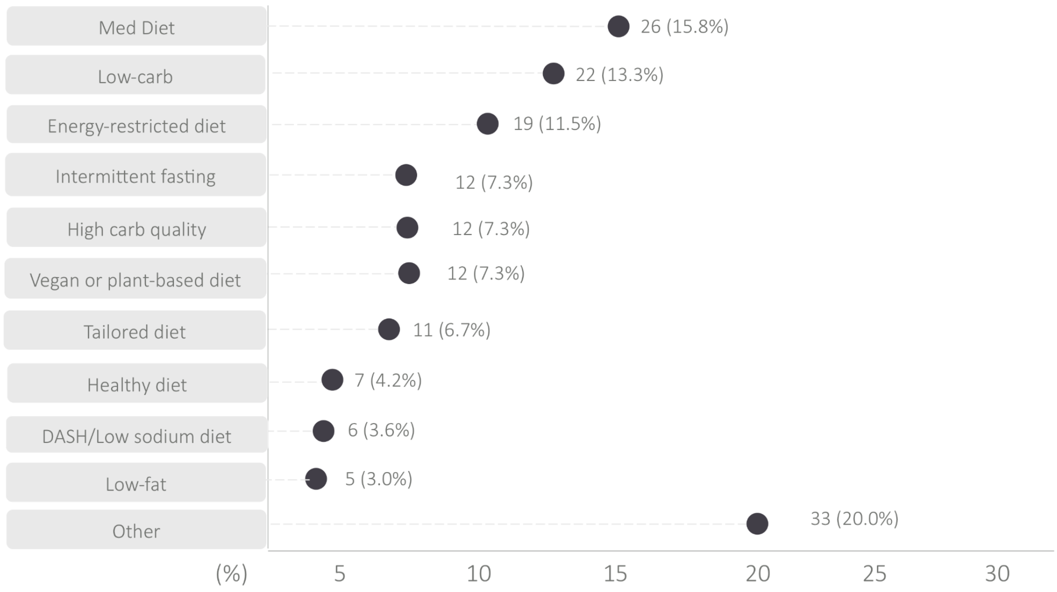


**Supplementary Figure 2**: Absolute and relative frequency of different types of supplements (A; n= 405) and complete diet or dietary pattern (B; n = 165) adopted as intervention by RCT protocols.

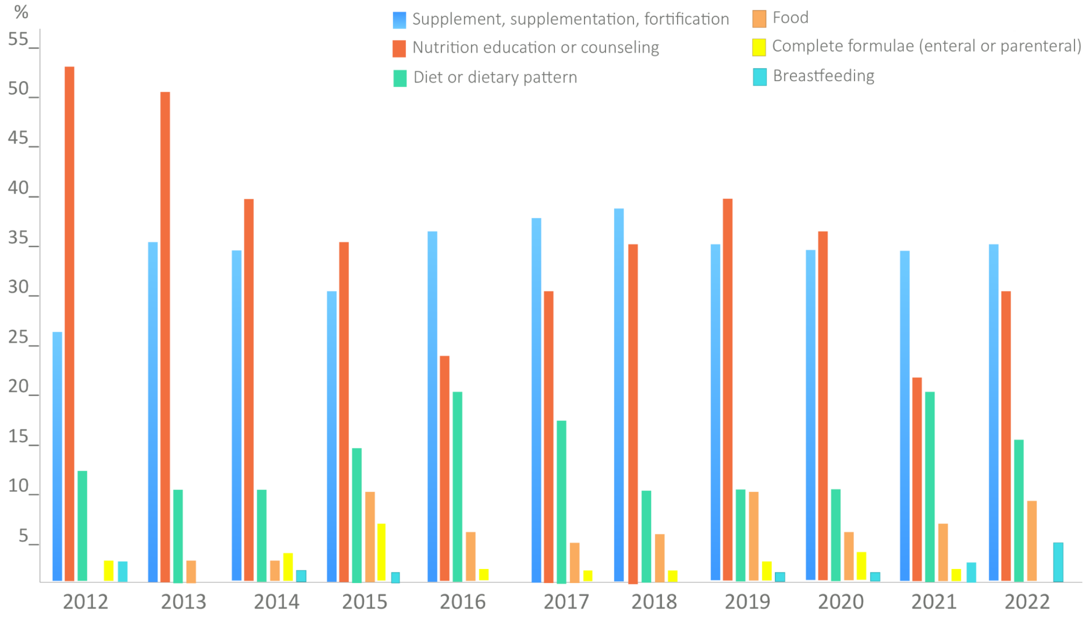


**Supplementary Figure 3**: Intervention categories according to the publication year of nutrition or diet-related RCT protocols.

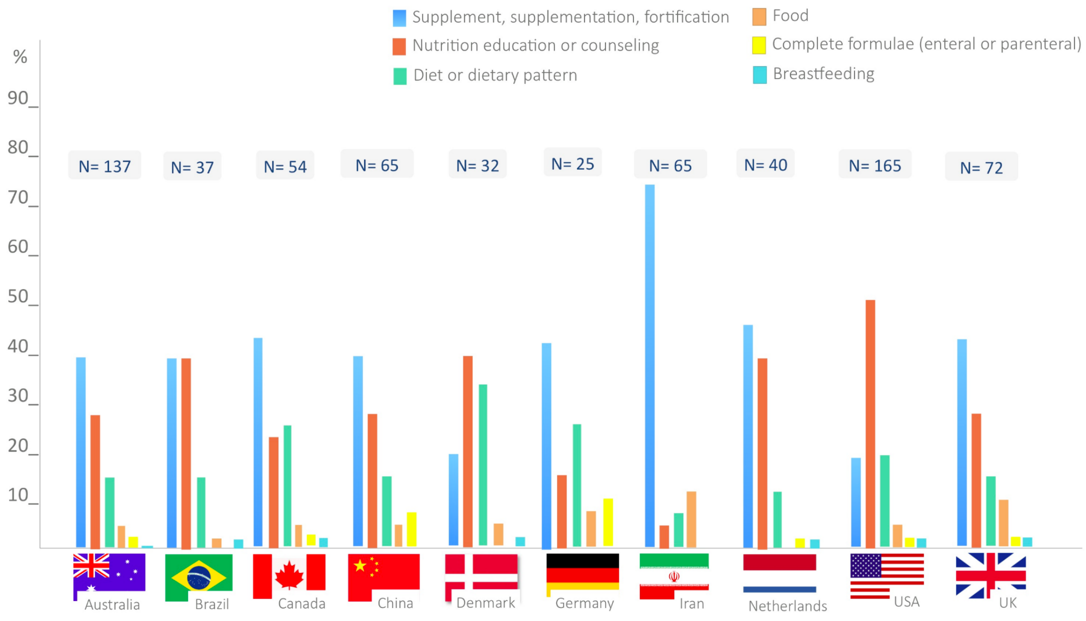


**Supplementary Figure 4**: Intervention's categories according to the country of nutrition or diet-related RCT protocols (sample of countries with at least 25 publications).

**Supplementary Table 1**. Relative frequency of protocols published in journals with an endorsement of reporting guidelines that cited them.

| Mentioned by the protocols | **Endorsement by the journals that published the protocols** | | | | | |
|---|---|---|---|---|---|---|
| | **SPIRIT** | | **CONSORT** | | **TIDIeR** | |
| | Yes | No | Yes | No | Yes | No |
| | 485 (45.4%) | 583 (54.6%) | 1016 (95.1%) | 52 (4.9%) | 7 (0.7%) | 1061 (99.3%) |
| Yes | 123 (25.4%) | 220 (37.7%) | 292 (28.7%) | 5 (9.6%) | 0 (0%) | 20 (1.9%) |
| No | 362 (74.6%) | 363 (62.3%) | 724 (71.3%) | 47 (90.4%) | 7 (100%) | 1041 (98.1%) |

**Supplementary Table 2.** Frequency of each component of the PICOS acronym for nutrition or diet related RCT protocols and practices of transparency and reproducibility stratified by the year of publication (n = 1068)

| | | **2012** | **2013** | **2014** | **2015** | **2016** | **2017** | **2018** | **2019** | **2020** | **2021** | **2022** |
|---|---|---|---|---|---|---|---|---|---|---|---|---|
| | N | 32 | 71 | 76 | 79 | 86 | 102 | 107 | 156 | 163 | 155 | 41 |
| P - Population (age-stage) | | | | | | | | | | | | |
| Infant | 66 (6.2%) | 2 (6.2%) | 2 (2.8%) | 5 (6.6%) | 8 (10.1%) | 5 (5.8%) | 8 (7.8%) | 5 (4.7%) | 8 (5.1%) | 8 (4.9%) | 9 (5.8%) | 6 (14.6%) |
| Children | 132 (12.4%) | 6 (18.8%) | 14 (19.7%) | 7 (9.2%) | 11 (13.9%) | 10 (11.6%) | 13 (12.7%) | 14 (13.1%) | 26 (16.7%) | 16 (9.8%) | 15 (9.7%) | 0 (0%) |
| Adolescent | 63 (5.9%) | 2 (6.2%) | 5 (7.0%) | 1 (1.3%) | 9 (11.4%) | 6 (7.0%) | 9 (8.8%) | 5 (4.7%) | 9 (5.8%) | 10 (6.1%) | 6 (3.9%) | 1 (2.4%) |
| Adult | 251 (23.5%) | 7 (21.9%) | 16 (22.5%) | 15 (19.7%) | 21 (26.6%) | 20 (23.3%) | 30 (29.4%) | 22 (20.6%) | 40 (25.6%) | 40 (24.5%) | 32 (20.6%) | 8 (19.5%) |
| Adult and Elderly | 350 (32.8%) | 5 (15.6%) | 22 (31.0%) | 30 (39.5%) | 22 (27.8%) | 26 (30.2%) | 30 (29.4%) | 37 (34.6%) | 48 (30.8%) | 56 (34.4%) | 61 (39.4%) | 13 (31.7%) |
| Elderly | 76 (7.1%) | 4 (12.5%) | 4 (5.6%) | 5 (6.6%) | 4 (5.1%) | 4 (4.7%) | 2 (2.%) | 11 (10.3%) | 11 (7.1%) | 14 (8.6%) | 14 (9.0%) | 3 (7.3%) |

| | | 2012 | 2013 | 2014 | 2015 | 2016 | 2017 | 2018 | 2019 | 2020 | 2021 | 2022 |
|---|---|---|---|---|---|---|---|---|---|---|---|---|
| Pregnant | 99 (9.3%) | 3 (9.4%) | 9 (12.7%) | 10 (13.2%) | 5 (6.3%) | 9 (10.5%) | 11 (10.8%) | 11 (10.3%) | 9 (5.8%) | 13 (8.0%) | 12 (7.7%) | 7 (17.1%) |
| Postmenopausal women | 9 (0.8%) | 0 (0%) | 0 (0%) | 1 (1.3%) | 0 (0%) | 4 (4.7%) | 0 (0%) | 0 (0%) | 0 (0%) | 1 (0.6%) | 3 (1.9%) | 0 (0%) |
| Families | 53 (5.0%) | 3 (9.4%) | 5 (7.0%) | 1 (1.3%) | 6 (7.6%) | 4 (4.7%) | 4 (3.9%) | 4 (3.7%) | 8 (5.1%) | 12 (7.4%) | 5 (3.2%) | 1 (2.4%) |
| Mother and infant | 19 (1.8%) | 0 (0%) | 2 (2.8%) | 2 (2.6%) | 2 (1.5%) | 1 (1.2%) | 1 (1.0%) | 3 (2.8%) | 2 (1.3%) | 2 (1.2%) | 3 (1.9%) | 1 (2.4%) |
| P - population (clinical condition) | | | | | | | | | | | | |
| None disease | 392 | 16 (50.0%) | 27 (38.0%) | 27 (35.5) | 26 (32.9%) | 34 (39.5%) | 43 (42.2%) | 38 (35.5%) | 67 (42.9%) | 42 (25.8%) | 59 (38.1%) | 13 (31.7%) |
| With a specific disease | 676 (63.3%) | 16 (50.0%) | 44 (62.0%) | 49 (64.5) | 53 (67.1%) | 52 (60.5%) | 59 (57.8%) | 69 (64.5%) | 89 (57.1%) | 121 (74.2%) | 96 (61.9%) | 28 (68.3%) |
| I - Intervention (type) | | | | | | | | | | | | |
| Isolated | 724 (67.8%) | 18 (56.2%) | 34 (47.9%) | 52 (68.4%) | 47 (59.5%) | 61 (70.9%) | 69 (67.6%) | 70 (65.4%) | 106 (67.9%) | 114 (69.9%) | 119 (76.8%) | 34 (82.9%) |
| Combined with exercise | 98 (9.2%) | 3 (9.4%) | 7 (9.9%) | 7 (9.2%) | 10 (12.7%) | 8 (9.3%) | 11 (10.8%) | 14 (13.1%) | 12 (7.7%) | 19 (11.7%) | 7 (4.5%) | 0 (0%) |

| | | 2012 | 2013 | 2014 | 2015 | 2016 | 2017 | 2018 | 2019 | 2020 | 2021 | 2022 |
|---|---|---|---|---|---|---|---|---|---|---|---|---|
| Combined with drug | 18 (1.7%) | 0 (0%) | 1 (1.4%) | 0 (0%) | 1 (1.3%) | 0 (0%) | 2 (2.0%) | 3 (2.8%) | 4 (2.6%) | 4 (2.5%) | 2 (1.3%) | 1 (2.4%) |
| Combined with other medical care | 18 (1.7%) | 0 (0%) | 1 (1.4%) | 0 (0%) | 2 (2.5%) | 2 (2.3%) | 0 (0%) | 3 (2.8%) | 3 (1.9%) | 3 (1.8%) | 4 (2.6%) | 0 (0%) |
| Part of lifestyle modification | 127 (11.9%) | 7 (21.9%) | 20 (28.2%) | 10 (13.2%) | 15 (19.0%) | 6 (7.0%) | 14 (13.7%) | 8 (7.5%) | 15 (9.6%) | 14 (8.6%) | 14 (9.0%) | 4 (9.8%) |
| Part of health program | 83 (7.8%) | 4 (12.5%) | 8 (11.3%) | 7 (9.2%) | 4 (5.1%) | 9 (10.5%) | 6 (5.9%) | 9 (8.4%) | 16 (10.3%) | 9 (5.5%) | 9 (5.8%) | 2 (4.9%) |
| I - Intervention (category) | | | | | | | | | | | | |
| Food | 78 (7.3%) | 0 (0%) | 2 (2.8%) | 3 (3.9%) | 7 (8.9%) | 6 (7.0%) | 6 (5.9%) | 8 (7.5%) | 17 (10.9%) | 12 (7.4%) | 13 (8.4%) | 4 (9.8%) |
| Diet or dietary pattern | 165 (15.4%) | 4 (12.5%) | 7 (9.9%) | 8 (10.5%) | 12 (15.2%) | 20 (23.3%) | 19 (18.6%) | 14 (13.1%) | 17 (10.9%) | 24 (14.7%) | 33 (21.3%) | 7 (17.1%) |
| Supplementation | 405 (37.9%) | 9 (28.1%) | 26 (36.6%) | 26 (34.2%) | 24 (30.4%) | 33 (38.4%) | 40 (39.2%) | 44 (41.1%) | 56 (35.9%) | 69 (42.3%) | 63 (40.6%) | 15 (36.6%) |
| Complete formulae | 37 (3.5%) | 1 (3.1%) | 0 (0%) | 4 (5.3%) | 6 (7.6%) | 2 (2.3%) | 3 (2.9%) | 2 (1.9%) | 6 (3.8%) | 9 (5.5%) | 4 (2.6%) | 0 (0%) |
| Nutrition Education | 354 (33.1%) | 17 (53.1%) | 36 (50.7%) | 31 (40.8%) | 28 (35.4%) | 21 (24.4%) | 32 (31.4%) | 39 (36.4%) | 57 (36.5%) | 46 (28.2%) | 35 (22.6%) | 12 (29.3%) |

| | | 2012 | 2013 | 2014 | 2015 | 2016 | 2017 | 2018 | 2019 | 2020 | 2021 | 2022 |
|---|---|---|---|---|---|---|---|---|---|---|---|---|
| Breastfeeding | 17 (1.6%) | 1 (3.1%) | 0 (0%) | 2 (2.6%) | 1 (1.3%) | 0 (0%) | 0 (0%) | 0 (0%) | 2 (1.3%) | 2 (1.2%) | 6 (3.9%) | 3 (7.3%) |
| Other | 12 (1.1%) | 0 (0%) | 0 (0%) | 2 (2.6%) | 1 (1.3%) | 4 (4.7%) | 2 (2.0%) | 0 (0%) | 1 (0.6%) | 1 (0.6%) | 1 (0.6%) | 0 (0%) |
| C - Comparator | | | | | | | | | | | | |
| Placebo | 362 (33.9%) | 7 (21.9%) | 26 (36.6%) | 23 (30.3%) | 26 (32.9%) | 29 (33.7%) | 36 (35.3%) | 30 (28.0%) | 55 (35.3%) | 58 (35.6%) | 58 (37.4%) | 14 (34.1%) |
| Other Intervention | 289 (27.1%) | 12 (37.5%) | 14 (19.7%) | 20 (26.3%) | 23 (29.1%) | 24 (27.9%) | 31 (30.4%) | 32 (29.9%) | 36 (23.1%) | 48 (29.4%) | 40 (25.8%) | 9 (22.0%) |
| Usual care | 316 (29.6%) | 7 (21.9%) | 22 (31.0%) | 25 (32.9%) | 27 (34.2%) | 25 (29.1%) | 27 (26.5%) | 35 (32.7%) | 48 (30.8%) | 38 (23.3%) | 46 (29.7%) | 16 (39.0%) |
| No control | 101 (9.5%) | 6 (18.8%) | 9 (12.7%) | 8 (10.5%) | 3 (3.8%) | 8 (9.3%) | 8 (7.5%) | 10 (9.3%) | 17 (10.9%) | 19 (11.7%) | 11 (7.1%) | 2 (4.9%) |
| O - Primary outcome (three most frequent categories) | | | | | | | | | | | | |
| Clinical status | 308 (28.8%) | 6 (18.8%) | 22 (31.0%) | 21 (27.6%) | 19 (24.1%) | 27 (31.4%) | 32 (31.4%) | 22 (20.6%) | 45 (28.8%) | 45 (27.6%) | 58 (37.4%) | 11 (26.8%) |
| Nutritional Status | 247 (23.1%) | 15 (46.9%) | 25 (35.2%) | 12 (15.8%) | 18 (22.8%) | 23 (26.7%) | 23 (22.5%) | 23 (21.5%) | 36 (23.1%) | 36 (22.1%) | 27 (17.4%) | 9 (22.0%) |

| | | 2012 | 2013 | 2014 | 2015 | 2016 | 2017 | 2018 | 2019 | 2020 | 2021 | 2022 |
|---|---|---|---|---|---|---|---|---|---|---|---|---|
| Frequency or severity of disease | 238 (22.3%) | 5 (15.6%) | 17 (23.9%) | 17 (22.4%) | 22 (27.8%) | 18 (20.9%) | 21 (20.6%) | 20 (18.7%) | 31 (19.9%) | 42 (25.8%) | 35 (22.6%) | 10 (24.4%) |
| S - Study design | | | | | | | | | | | | |
| Crossover | 54 | 1 (3.1%) | 4 (5.6%) | 5 (6.6%) | 4 (5.1%) | 5 (5.8%) | 6 (5.9%) | 6 (5.6%) | 7 (4.5%) | 7 (4.3%) | 7 (4.5%) | 2 (4.9%) |
| Parallel | 1014 | 31 (96.9%) | 67 (94.4%) | 71 (93.4%) | 75 (94.9%) | 81 (94.2%) | 96 (94.1%) | 101 (94.4%) | 149 (95.5%) | 156 (95.7%) | 148 (95.5%) | 39 (95.5%) |
| Practices of transparency and reproducibility | | | | | | | | | | | | |
| Protocol register | 1006 (94.2%) | 30 (93.8%) | 61 (85.9%) | 72 (94.7%) | 72 (91.1%) | 80 (93.0%) | 100 (98.0%) | 102 (95.3%) | 147 (94.2%) | 156 (95.7%) | 147 (94.8%) | 39 (95.1%) |
| Conflicts of interest statement | 953 (89.2%) | 29 (90.6%) | 58 (81.7%) | 69 (90.8%) | 71 (89.9%) | 73 (84.9%) | 96 (94.1%) | 95 (88.8%) | 136 (87.2%) | 146 (89.6%) | 141 (91.0%) | 39 (95.1%) |
| Funding statement | 1043 (97.7%) | 32 (100%) | 68 (95.8%) | 73 (96.1%) | 76 (96.2%) | 82 (95.3%) | 100 (98.0%) | 105 (98.1%) | 154 (98.7%) | 160 (98.2%) | 152 (98.1%) | 41 (100%) |

**Supplementary Table 3.** Absolute and relative frequency of each component of the PICOS acronyms and other information related to transparency for nutrition or diet-related RCT protocols according to the five most frequent countries for trials conduction.

| **Characteristics** | **USA (n = 165)** | **UK (n = 72)** | **Iran (n = 65)** | **China (n = 65)** | **Australia (n=137)** |
|---|---|---|---|---|---|
| *P - population (life-stage)* | | | | | |
| Infant | 6 (3.6%) | 4 (5.6%) | 0 (0%) | 4 (6.2%) | 4 (2.9%) |
| Children | 32 (19.4%) | 4 (5.6%) | 2 (3.1%) | 6 (9.2%) | 16 (11.7%) |
| Adolescent | 11 (6.7%) | 4 (5.6%) | 3 (4.6%) | 1 (1.5%) | 7 (5.1%) |
| Adult | 37 (22.4%) | 20 (27.8%) | 31 (47.7%) | 14 (21.5%) | 35 (25.5%) |
| Adult and Elderly | 38 (23.0%) | 29 (40.3%) | 26 (40.0%) | 24 (36.9%) | 47 (34.3%) |
| Elderly | 6 (3.6%) | 8 (11.1%) | 0 (0%) | 3 (4.6%) | 12 (8.8%) |
| Pregnant | 16 (9.7%) | 2 (2.8%) | 2 (3.1%) | 6 (9.2%) | 14 (10.2%) |
| Postmenopausal women | 3 (1.8%) | 0 (0%) | 1 (1.5%) | 3 (4.6%) | 1 (0.7%) |
| Families | 33 (20.0%) | 1 (1.4%) | 0 (0%) | 4 (6.2%) | 2 (1.5%) |
| Mother and infant | 3 (1.8%) | 1 (1.4%) | 0 (0%) | 1 (1.5%) | 0 (0%) |
| *P - population (clinical condition)* | | | | | |
| No disease | 63 (38.2%) | 22 (30.6%) | 7 (10.8%) | 18 (27.7%) | 48 (35.0%) |
| Specific disease | 102 (61.8%) | 50 (69.4%) | 58 (89.2%) | 47 (72.3%) | 89 (65.0%) |

| Characteristics | USA (n = 165) | UK (n = 72) | Iran (n = 65) | China (n = 65) | Australia (n=137) |
|---|---|---|---|---|---|
| *I - Intervention (type of intervention)* | | | | | |
| Isolated | 89 (53.9%) | 48 (66.7%) | 61 (93.8%) | 48 (73.8%) | 95 (69.3%) |
| Combined with exercise | 19 (11.5%) | 9 (12.5%) | 1 (1.5%) | 8 (12.3%) | 14 (10.2%) |
| Combined with drug | 0 (0%) | 4 (5.6%) | 0 (0%) | 3 (4.6%) | 0 (0%) |
| Combined with other medical care | 1 (0.6%) | 1 (1.4%) | 1 (1.5%) | 0 (0%) | 2 (1.5%) |
| Part of lifestyle modification | 34 (20.6%) | 7 (9.7%) | 1 (1.5%) | 2 (3.1%) | 18 (13.1%) |
| Part of health program | 22 (13.3%) | 3 (4.2%) | 1 (1.5%) | 4 (6.2%) | 8 (5.8%) |
| *I - Intervention (categories of intervention)* | | | | | |
| Food | 10 (6.1%) | 8 (11.1%) | 8 (12.3%) | 3 (4.6%) | 7 (5.1%) |
| Diet or dietary pattern | 34 (20.6%) | 11 (15.3%) | 5 (7.7%) | 11 (16.9%) | 22 (16.1%) |
| Supplementation | 30 (18.2%) | 31 (43.1%) | 49 (75.5%) | 25 (38.5%) | 53 (38.7%) |
| Complete formulae enteral or parenteral | 2 (1.2%) | 1 (1.4%) | 0 (0%) | 6 (9.2%) | 3 (2.2%) |
| Nutrition Education | 84 (50.9%) | 20 (27.8%) | 3 (4.6%) | 18 (27.7%) | 49 (35.8%) |
| Breastfeeding | 3 (1.8%) | 1 (1.4%) | 0 (0%) | 1 (1.5%) | 1 (0.7%) |
| Other | 0 (0%) | 0 (0%) | 0 (0%) | 0 (0%) | 2 (1.5%) |

| **Characteristics** | **USA (n = 165)** | **UK (n = 72)** | **Iran (n = 65)** | **China (n = 65)** | **Australia (n=137)** |
|---|---|---|---|---|---|
| *C - Comparator* | | | | | |
| Placebo | 33 (20.0%) | 27 (37.5%) | 48 (73.8%) | 23 (35.4%) | 48 (35.0%) |
| Other Intervention | 63 (38.2%) | 20 (27.8%) | 9 (13.8%) | 15 (23.1%) | 26 (19.0%) |
| Usual care | 52 (31.5%) | 18 (25.0%) | 5 (7.7%) | 18 (27.7%) | 50 (36.5%) |
| No control | 17 (10.3%) | 7 (9.7%) | 3 (4.6%) | 9 (13.8%) | 13 (9.5%) |
| *O - Primary Outcomes ( most frequent)* | | | | | |
| Clinical status | 36 (21.8%) | 14 (19.4%) | 49 (75.4%) | 20 (30.8%) | 33 (24.1%) |
| Nutritional Status | 62 (37.6%) | 11 (15.3%) | 7 (10.8%) | 13 (20.0%) | 25 (18.2%) |
| Frequency or severity of disease | 22 (13.3%) | 15 (20.8%) | 14 (21.5%) | 20 (30.8%) | 32 (23.4%) |
| Withdrawal, drop-out or adherence | 13 (7.9%) | 16 (22.2%) | 3 (4.6%) | 1 (1.5%) | 8 (5.8%) |
| *S - Study design* | | | | | |
| Crossover | 9 (5.5%) | 5 (6.9%) | 2 (3.1%) | 0 (0%) | 7 (5.1%) |
| Parallel | 156 (94.5%) | 67 (93.1%) | 63 (96.9%) | 65 (100%) | 130 (94.9%) |

| **Characteristics** | **USA (n = 165)** | **UK (n = 72)** | **Iran (n = 65)** | **China (n = 65)** | **Australia (n=137)** |
|---|---|---|---|---|---|
| *Transparency and reproducibility practices* | | | | | |
| Protocol register | 137 (83.0%) | 65 (90.3%) | 64 (98.5%) | 64 (98.5%) | 132 (96.4%) |
| Conflicts of interest statement | 108 (66.4%) | 68 (94.4%) | 64 (98.5%) | 63 (96.9%) | 130 (94.9%) |
| Funding statement | 161 (97.6%) | 71 (98.6%) | 62 (95.4%) | 62 (95.4%) | 130 (94.9%) |
| Mention of SPIRIT | 23 (13.9%) | 22 (30.6%) | 36 (55.4%) | 23 (35.4%) | 34 (24.8%) |
| Mention of TIDIeR | 1 (0.6%) | 6 (8.3%) | 0 (0%) | 0 (0%) | 3 (2.2%) |
| Mention of CONSORT | 35 (21.2%) | 20 (27.8%) | 13 (20.0%) | 15 (23.1%) | 56 (40.9%) |

**Supplementary Table 4.** Absolute and relative frequency of each component of the PICOS acronyms and other information related to transparency for nutrition or diet-related RCT protocols focusing on patients with cancer and cardiovascular disease

| | Cancer (n = 76) | | Cardiovascular disease (n = 90) | |
|---|---|---|---|---|
| | n | % | n | % |
| **Population categories** | | | | |
| Infant | 0 | 0 | 1 | 1.1 |
| Children | 2 | 2.6 | 1 | 1.1 |
| Adolescent | 2 | 2.6 | 2 | 2.2 |
| Adult | 6 | 7.9 | 7 | 7.8 |
| Adult and Elderly | 61 | 80.3 | 73 | 81.1 |
| Elderly | 1 | 1.3 | 7 | 7.8 |
| Pregnant | 0 | 0 | 1 | 1.1 |
| Postmenopausal women | 0 | 0 | 1 | 1.1 |
| Families | 3 | 3.9 | 1 | 1.1 |
| Mother and infant | 0 | 0 | 0 | 0 |
| **Intervention type** | | | | |
| Isolated | 48 | 63.2 | 67 | 74.4 |
| Combined with exercise | 16 | 21.1 | 5 | 5.6 |
| Combined with drug | 3 | 3.9 | 1 | 1.1 |

| | | | | |
|---|---|---|---|---|
| Combined with other medical care | 0 | 0 | 0 | 0 |
| Part of lifestyle modification | 8 | 10.5 | 7 | 7.8 |
| Part of health program | 1 | 1.3 | 10 | 11.1 |
| **Intervention categories** | | | | |
| Food | 3 | 3.9 | 12 | 13.3 |
| Diet or dietary pattern | 22 | 28.9 | 19 | 21.1 |
| Supplementation | 25 | 32.9 | 33 | 36.7 |
| Complete formulae | 11 | 14.5 | 2 | 2.2 |
| Nutrition Education | 15 | 19.7 | 24 | 26.7 |
| Breastfeeding | 0 | 0 | 0 | 0 |
| Other | 0 | 0 | 0 | 0 |
| **Comparator categories** | | | | |
| Placebo | 20 | 26.3 | 34 | 37.8 |
| Other Intervention | 21 | 27.6 | 29 | 32.2 |
| Usual care | 32 | 42.1 | 23 | 25.6 |
| No control | 3 | 3.9 | 4 | 4.4 |
| **Primary outcomes categories (the three most frequent)** | | | | |
| Clinical status | 11 | 14.5 | 39 | 43.3 |
| Nutritional Status | 15 | 19.7 | 7 | 7.8 |
| Frequency or severity of disease | 19 | 25.0 | 22 | 24.4 |
| **Study design** | | | | |

| | | | | |
|---|---|---|---|---|
| Crossover | 1 | 1.3 | 5 | 5.6 |
| Parallel | 75 | 98.7 | 85 | 94.4 |
| **Other information - disclosure of conflicts of interest** | | | | |
| No reporting | 7 | 9.2 | 12 | 13.3 |
| Reporting of conflicts of interest | 11 | 14.5 | 13 | 14.4 |
| Reporting of none conflicts of interest | 58 | 76.3 | 65 | 72.2 |
| **Other information - disclosure of funding** | | | | |
| No reporting | 1 | 1.3 | 2 | 2.2 |
| Reporting of funding | 67 | 88.2 | 87 | 96.7 |
| Reporting of none funding | 8 | 10.5 | 1 | 1.1 |
| **Other information - registry of protocol** | | | | |
| Yes | 69 | 90.8 | 88 | 97.8 |
| No | 7 | 9.2 | 2 | 2.2 |
